# Clinical implementation of RNA sequencing for Mendelian disease diagnostics

**DOI:** 10.1101/2021.04.01.21254633

**Authors:** Vicente A. Yépez, Mirjana Gusic, Robert Kopajtich, Christian Mertes, Nicholas H. Smith, Charlotte L. Alston, Rui Ban, Skadi Beblo, Riccardo Berutti, Holger Blessing, Elżbieta Ciara, Felix Distelmaier, Peter Freisinger, Johannes Häberle, Susan J. Hayflick, Maja Hempel, Yulia S. Itkis, Yoshihito Kishita, Thomas Klopstock, Tatiana D. Krylova, Costanza Lamperti, Dominic Lenz, Christine C. Makowski, Signe Mosegaard, Michaela F. Müller, Gerard Muñoz-Pujol, Agnieszka Nadel, Akira Ohtake, Yasushi Okazaki, Elena Procopio, Thomas Schwarzmayr, Joél Smet, Christian Staufner, Sarah L. Stenton, Tim M. Strom, Caterina Terrile, Frederic Tort, Rudy Van Coster, Arnaud Vanlander, Matias Wagner, Manting Xu, Fang Fang, Daniele Ghezzi, Johannes A. Mayr, Dorota Piekutowska-Abramczuk, Antonia Ribes, Agnès Rötig, Robert W. Taylor, Saskia B. Wortmann, Kei Murayama, Thomas Meitinger, Julien Gagneur, Holger Prokisch

## Abstract

**Background:** Lack of functional evidence hampers variant interpretation, leaving a large proportion of cases with a suspected Mendelian disorder without genetic diagnosis after genome or whole exome sequencing (WES). Research studies advocate to further sequence transcriptomes to directly and systematically probe gene expression defects. However, collection of additional biopsies, and establishment of lab workflows, analytical pipelines, and defined concepts in clinical interpretation of aberrant gene expression are still needed for adopting RNA-sequencing (RNA-seq) in routine diagnostics.

**Methods:** We implemented an automated RNA-seq protocol and a computational workflow with which we analyzed skin fibroblasts of 303 individuals with a suspected mitochondrial disease which previously underwent WES.

**Results:** We detected on average 12,500 genes per sample including around 60% disease genes - a coverage substantially higher than with whole blood, supporting the use of skin biopsies. We prioritized genes demonstrating aberrant expression, aberrant splicing, or mono-allelic expression. The pipeline required less than one week from sample preparation to result reporting and provided a median of eight disease-associated genes per patient for inspection. A genetic diagnosis was established for 16% of the 205 WES-inconclusive cases. Detection of aberrant expression was a major contributor to diagnosis including instances of 50% reduction, which, together with mono-allelic expression, allowed for the diagnosis of dominant disorders caused by haploinsufficiency. Moreover, calling aberrant splicing and variants from RNA-seq data enabled detecting and validating splice-disrupting variants, of which the majority fell outside WES-covered regions.

**Conclusion:** Together, these results show that streamlined experimental and computational processes can accelerate the implementation of RNA-seq in routine diagnostics.

**One sentence summary:** Implementation of RNA-seq as a complementary tool in standard diagnostics achieves a 16% in diagnosis rate over whole exome sequencing.

## Background

It is estimated that at least 3.5-6% of the human population is affected by a rare disease (1), of which presumably ∼80% have a genetic cause (2). Although not necessarily providing the cure, establishing the correct and timely diagnosis of a Mendelian disease can improve disease management, provide prognostic information, and inform genetic counselling (3-5). Clinical implementation of next generation sequencing (NGS), especially whole exome sequencing (WES), revolutionized genetic diagnostics of individuals suspected of having a Medelian disorder by improving diagnostic yield and accelerating discovery of novel disease genes (6,7). Nevertheless, the diagnostic yield of WES analysis rarely exceeds 50% and hence leaves the majority of patients without a genetic diagnosis (8-12). Inconclusive WES can be partially attributed to the challenges concerning variant detection, prioritization, and interpretation. Although whole genome sequencing (WGS) allows, in principle, the detection of all genomic variants, its clinical implementation has reported similar diagnostic rates to those of WES (13,14). This indicates that variant prioritization and interpretation are the main challenges in genetic diagnostics (15).

So far, variants predicted to have potentially large effects on protein function are limited to large copy number variations, loss-of-function variants such as frameshift, start loss, stop gain, and stop loss, and variants altering splice acceptor or donor dinucleotides (16). However, it has been suggested that up to 30% of pathogenic variants fall within non-coding regions (17,18). Moreover, multiplex splicing assays showed that splicing-disturbing variants include about 10% of pathogenic exonic variants and are difficult to predict (19,20). Although many *in silico* tools have been developed to predict the effect of a variant on transcription, splicing, or RNA stability, their accuracy remains too low to establish a firm diagnosis. Without the necessary functional validation using either a minigene or patient biopsy material, splice-region and non-coding variants remain variants of uncertain significance (VUS, (21)).

By directly probing transcript abundance and sequence on a transcriptome-wide basis, RNA-seq allows systematic identification of aberrant transcript events, defined as genes expressed at aberrant levels, aberrantly spliced genes, and mono-allelically expressed (MAE) rare variants. Detection of such events enables validation of VUS potentially affecting the transcript, re-interpretation of VUS when linked to an aberrant transcript event, and discovery of pathogenic variants not covered by WES. A recent study concluded that up to 31% splicing VUSs could reach either a likely pathogenic or likely benign classification from RNA-seq analysis (22). The application of RNA-seq has increased diagnostic rates by 8-36% across a variety of rare disorders and selected cohorts of up to approximately one hundred affected individuals (23-28). Besides increasing the diagnostic yield, RNA-seq can improve the understanding of the molecular pathomechanism of the variant and basic genetic mechanisms. This can also be of therapeutic importance, for instance, to design splicing-targeted treatments, as has been demonstrated for example in Duchenne muscular dystrophy (29). While these initial studies are promising, routine clinical implementation of RNA-seq requires robust and efficient computational workflows, establishment of quality controls, and adequate RNA source material.

Here, we report on our experience on the implementation of RNA-seq into clinical diagnostics using patient-derived skin fibroblasts (Fig. 1). We demonstrate the application of our validated computational workflow, DROP (30), which integrates preprocessing and quality control steps, as well as modules for detecting aberrant expression, aberrant splicing, and MAE (Fig. 1), and to which we have added a new module for RNA-seq based variant calling. We apply this workflow to a compendium of WES and RNA-seq samples of 303 individuals suspected of having a mitochondrial or other Mendelian disease, the largest such dataset to date. For each type of aberrant event, we examine their genetic background and provide diagnostic guidance through case studies. While our analysis is based solely on fibroblast-derived material and with the majority of individuals suspected of having a mitochondrial disorder, our study also addresses the value of other clinically-accessible tissues for the diagnosis of Mendelian disorders.

**Fig. 1.**
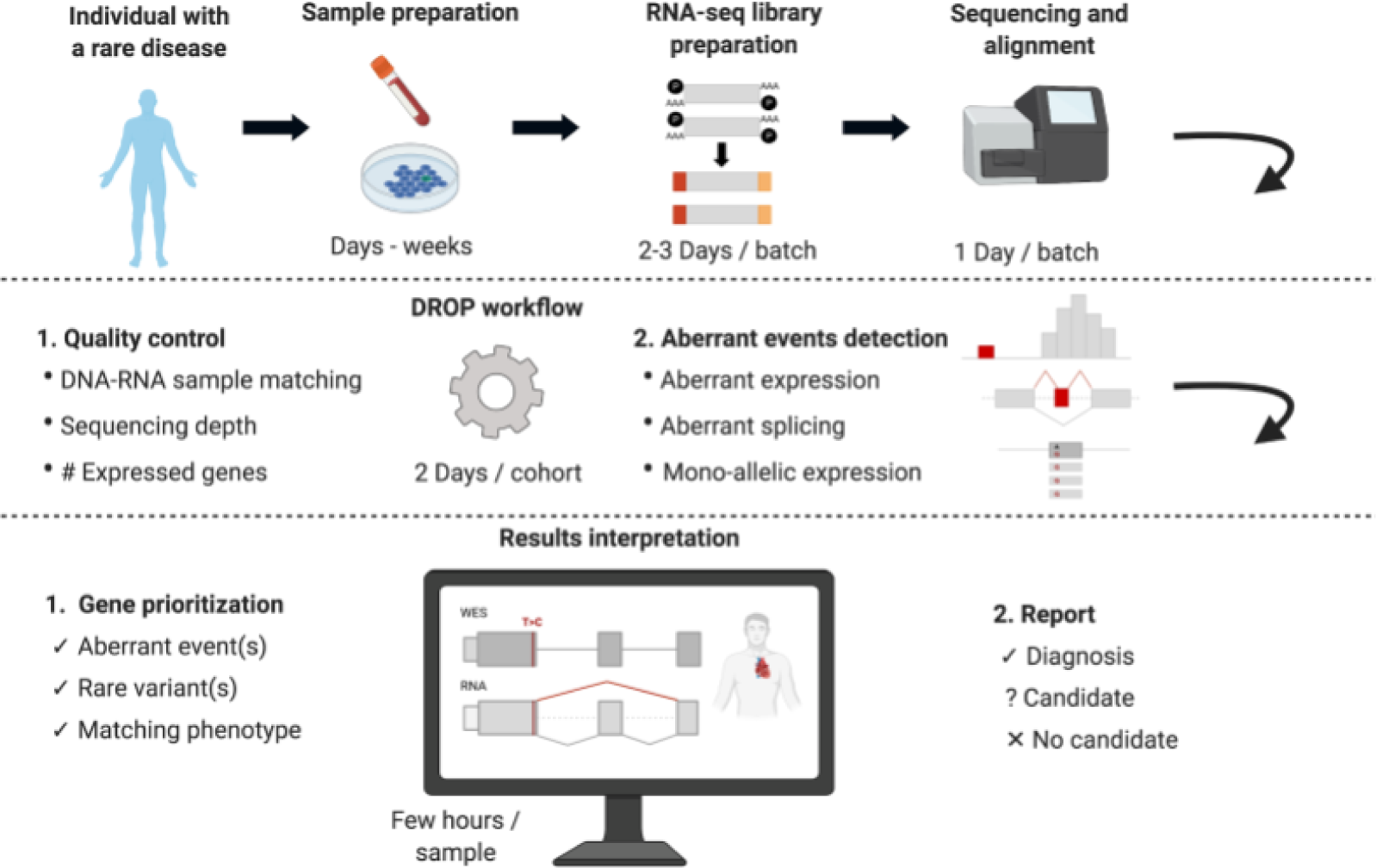
Experimental design of an RNA-seq based diagnostic study. First, individuals suspected of a Mendelian disorder are recruited for DNA sequencing. In addition, patient biopsy material is collected during the routine medical examination and prepared for RNA extraction. The sample preparation process can take from hours for biopsies to weeks for establishing a cell culture. RNA sequencing is then performed followed by alignment and quality control. The generated data go through DROP which consists of quality control steps and detection of aberrant RNA expression events. The results are then interpreted by sample, including association of aberrant RNA expression events with rare variant(s) and the function of affected genes with the patient phenotype, which can lead to new diagnoses or candidates. Experience-based estimated durations are provided for each step.

## Results

### RNA-seq analysis workflow

Extending the study of Kremer and colleagues (23) to support routine diagnostic testing, we recruited 303 individuals suspected to be affected by a Mendelian disorder with fibroblasts cell lines available within an international collaboration, and performed WES and RNA-seq on them (table S1, fig. S1, Materials and Methods). Almost all individuals (87%, 263 out of 303) were clinically suspected to suffer from a mitochondrial disease, presenting with a broad spectrum of clinical signs and symptoms. Mitochondrial disease represents an attractive class of rare disorders for the development and testing of systematic large-scale diagnostic screening approaches on account of significant clinical and genetic heterogeneity, with pathogenic variants described in more than 340 genes (31). The study cohort consists of 98 WES diagnosed cases used to establish a reference dataset of gene expression (some of which have been published as single gene studies (4,32–53) and 205 cases that remained inconclusive after WES (fig. S1). A single RNA-seq assay was performed per individual. A total of 101 samples were sequenced following a non-strand specific protocol and 202 following a strand-specific one using automated protocols minimizing sample handling and allowing highly reproducible results (Materials and Methods). We provide the gene expression count matrices, as well as the privacy-preserving count matrices of split and unsplit reads overlapping annotated splice sites, which can then be integrated by external users through DROP (Data availability).

**Table 1.**
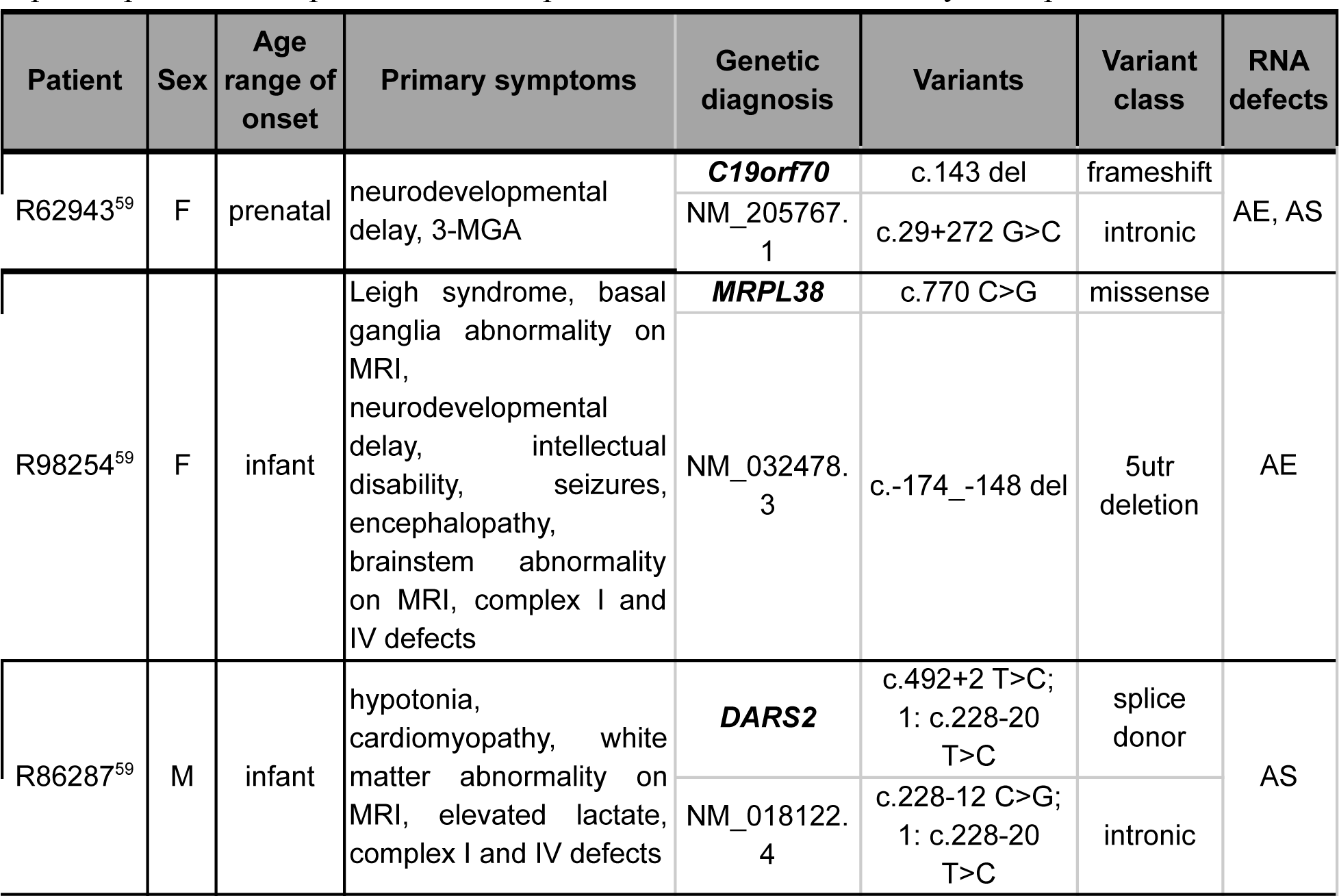
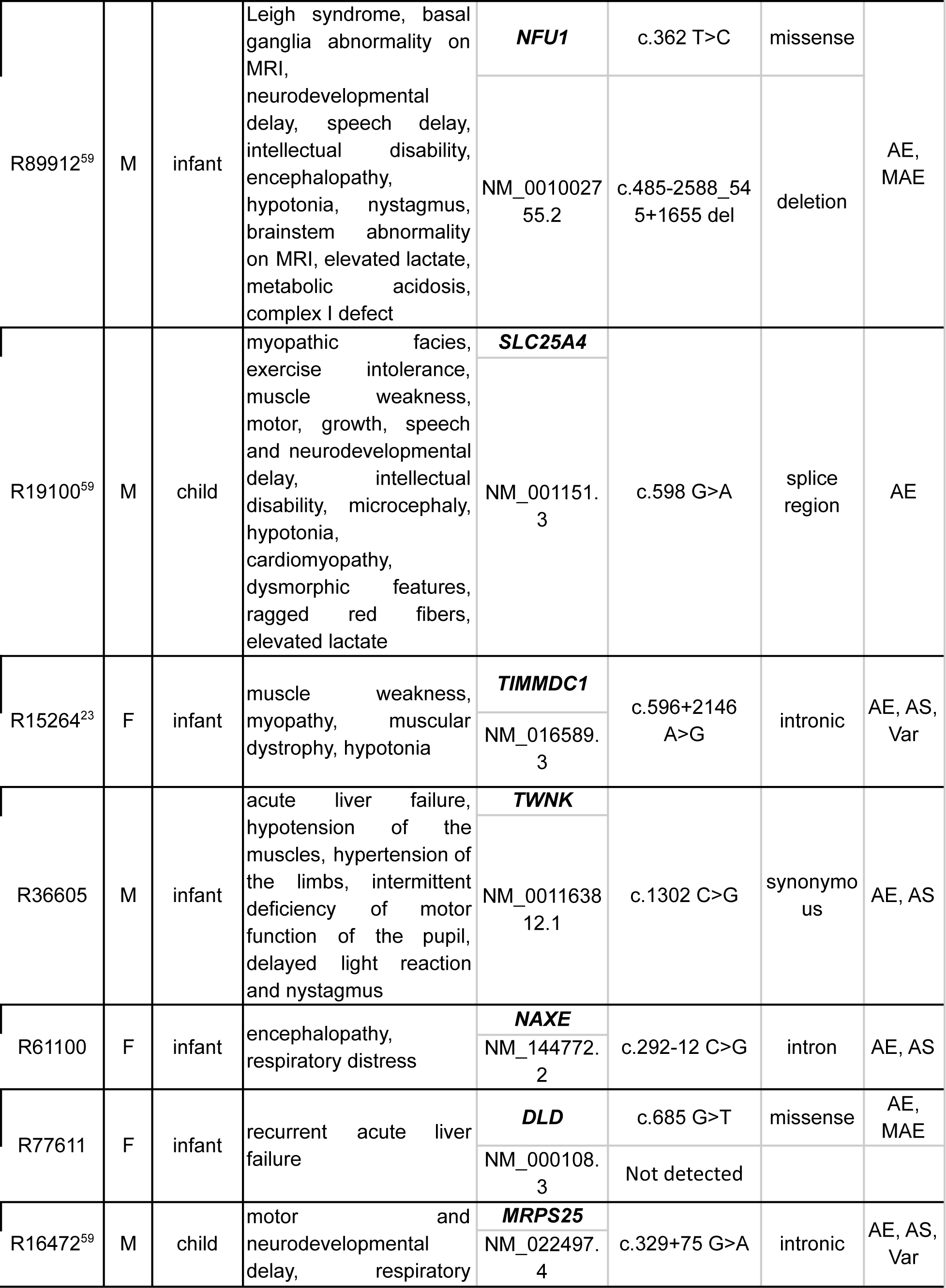
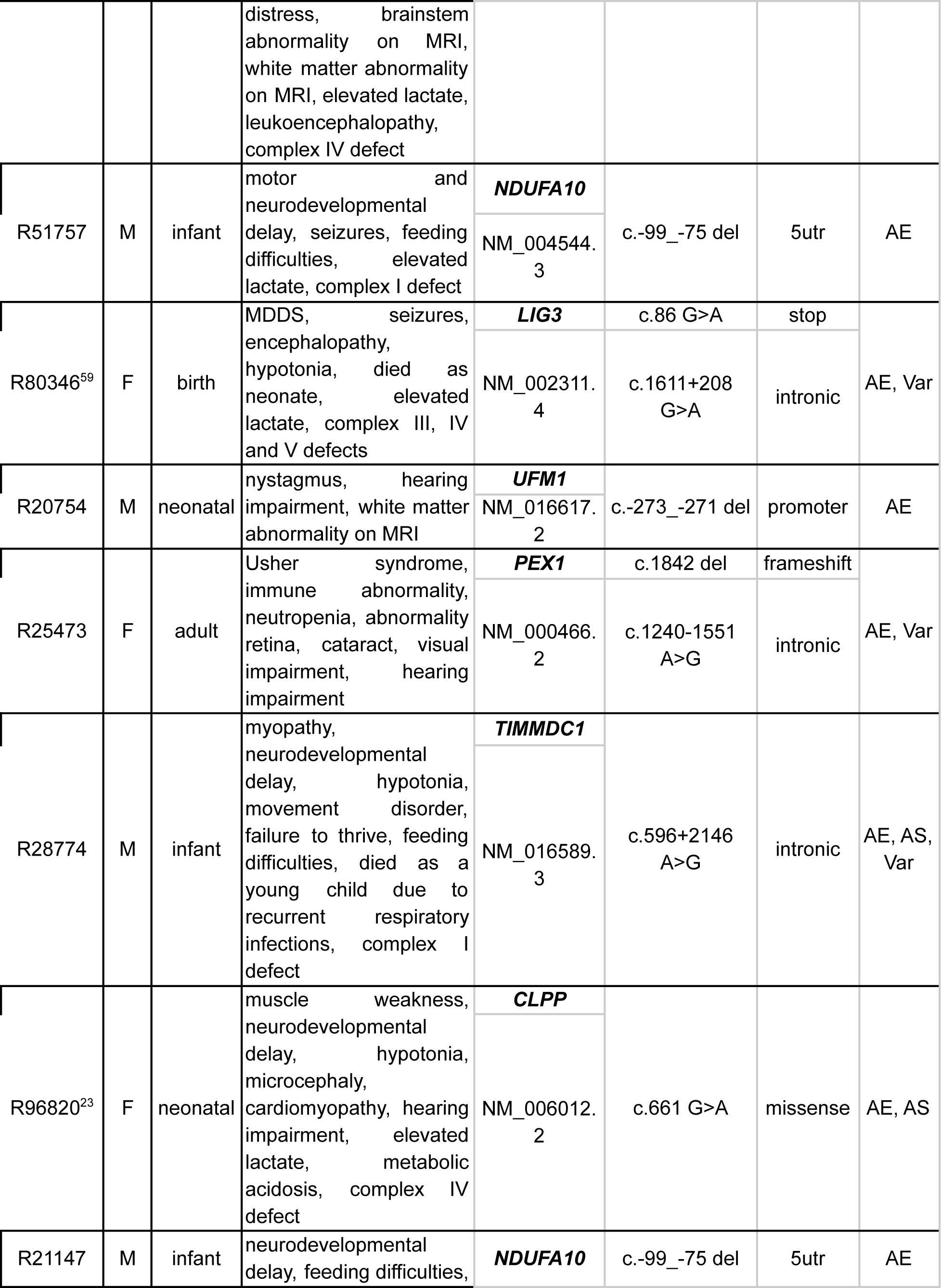
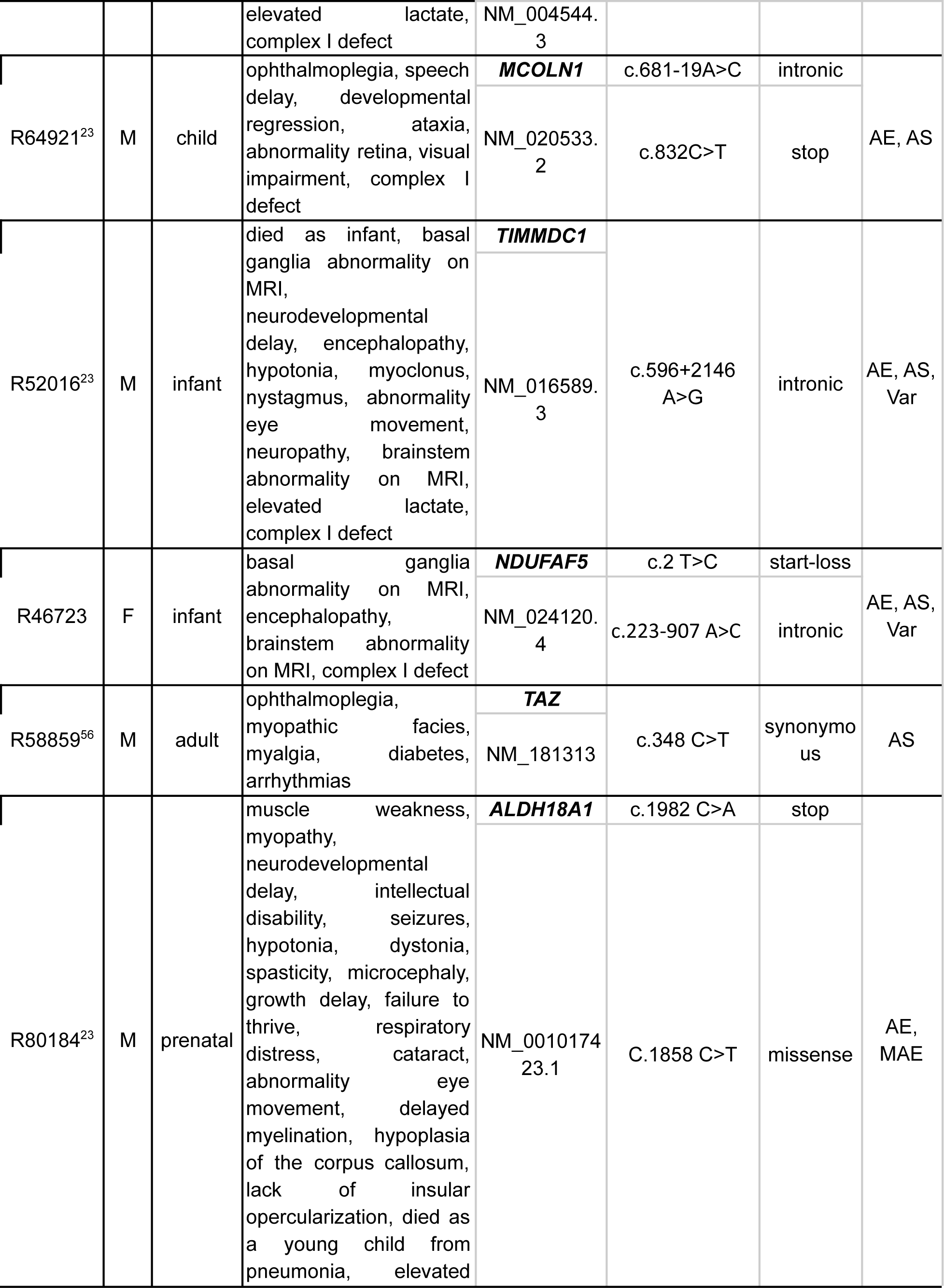
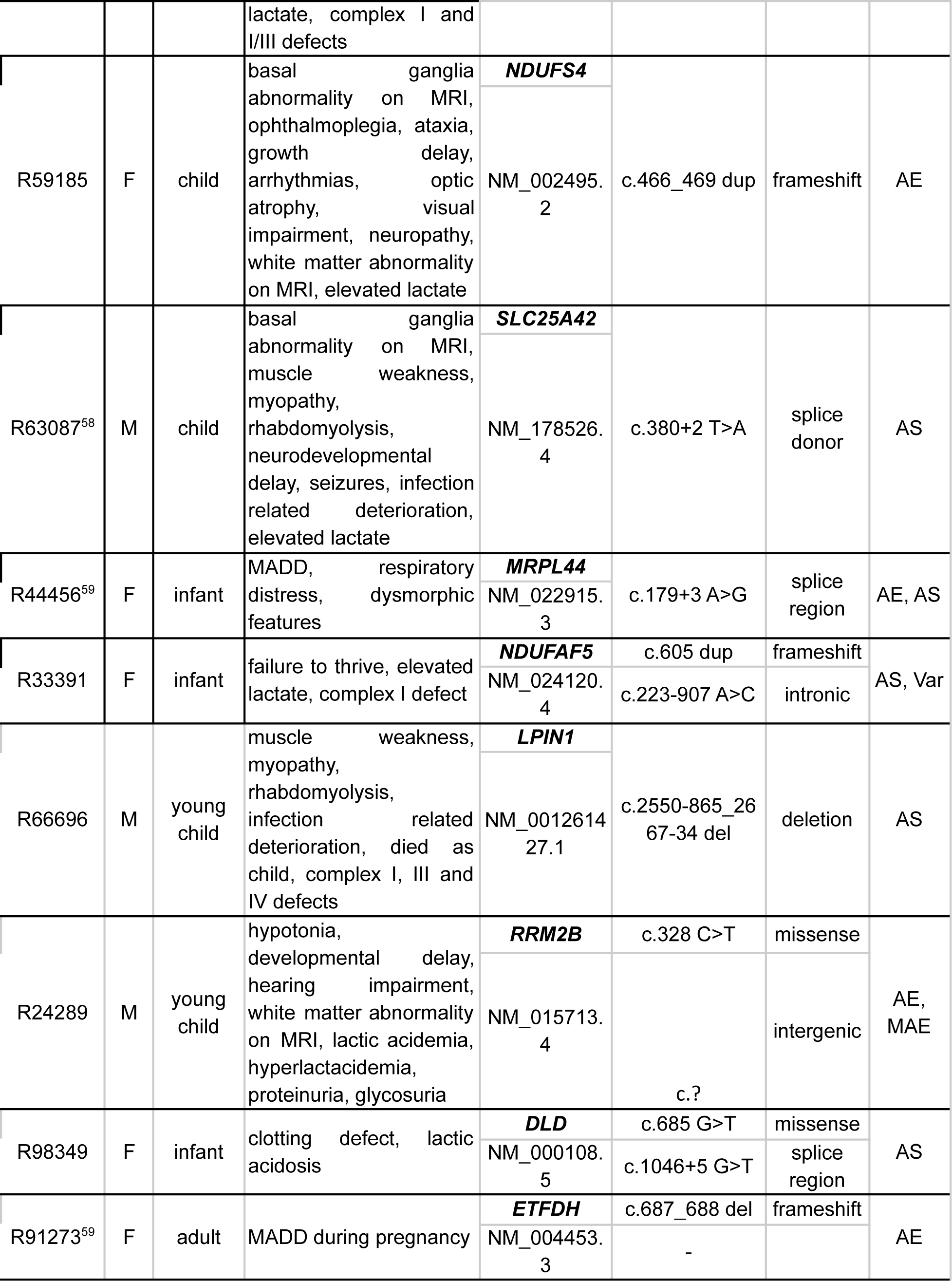
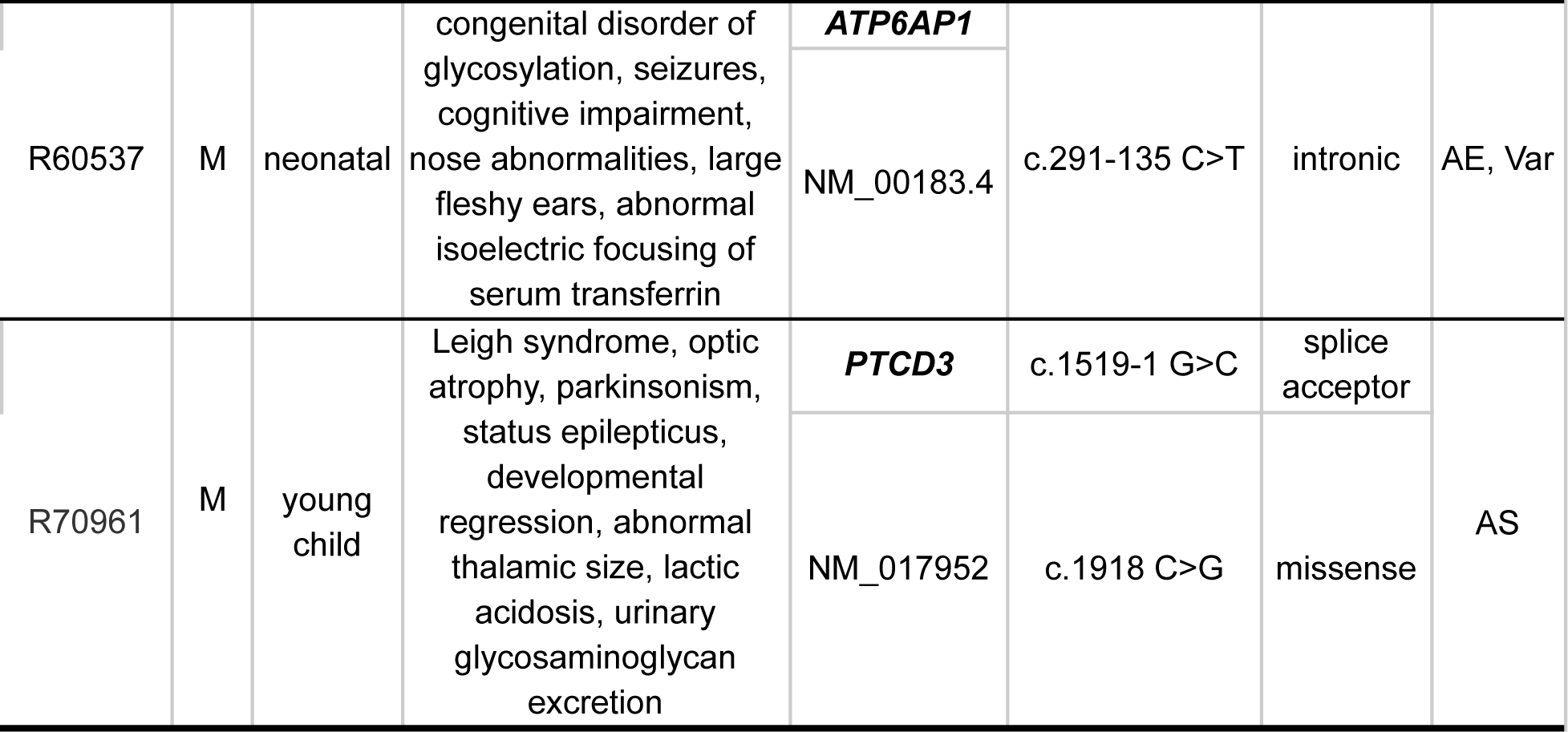
Summary of cases diagnosed via RNA-seq. AE: aberrant expression, AS: aberrant splicing, MAE: mono-allelic expression, Var: intronic variant detected via RNA-seq. The superscripts in certain patient ids correspond to the studies where they were published.

After alignment, RNA-seq data were analyzed using the computational workflow DROP (30), which ensures reproducibility, robustness, and scalability (Fig. 1, Materials and Methods). Quality control metrics were computed including the sequencing depth and percentage of high-quality reads aligned (using RNA-SeQC (54)), and the number of expressed genes per sample using DROP (fig. S2). DROP also computes the percentage of matching DNA-RNA variants to control for sample mismatches, which allowed us to reassign ten RNA-seq samples to their corresponding DNA (fig. S3, Materials and Methods). Afterward, DROP calls aberrant expression, aberrant splicing, and MAE using the statistical methods OUTRIDER (55), FRASER (56), and a negative binomial test (23), respectively. This yielded a median of 29 aberrant genes per sample, including eight where variants have been reported to cause a Mendelian disease in humans (OMIM (57), fig. S4). Aberrant events involving known disease-associated genes were then inspected manually in a case-by-case fashion comparing patient phenotype information with the phenotypes and mode of inheritance associated with the disease-associated gene (Fig. 2). For plausible candidate genes, we next inspected the sequencing data and searched for causative variants called by either WES or RNA-seq, and in some cases performed WGS followed by segregation analysis of the likely pathogenic variants (Fig. 2). This procedure led to a genetic diagnosis of 32 unsolved cases, representing 16% (95%CI: 11%-22%) of the WES-inconclusive cohort (Table 1, fig. S1), seven of which were previously published (23,58), and ten described in a companion manuscript (59). Among the 46 causative variants in these 32 cases, 13 (28%) were already classified as pathogenic or likely pathogenic, 10 (22%) required functional validation, 11 (24%) were not prioritized during WES-analysis, and 12 (26%) were not captured by WES. Three (25%) of the uncaptured group required WGS to identify the causative variant, while the other nine (75%) were detected using variant calling from RNA-seq (Table 1). In addition to the solved cases, we identified potential candidates in 12 cases: in 8 we identified a likely pathogenic change at the transcript level but have been unable to pinpoint the causative variant, and in 4 cases we identified aberrant expression and likely deleterious variants in candidate genes, representing likely novel disease genes (table S2, Fig. 2, fig. S1, fig. S4). These candidate cases are currently being investigated in follow-up studies. Here, we outline each screening step for the detection of aberrant events.

**Fig. 2.**
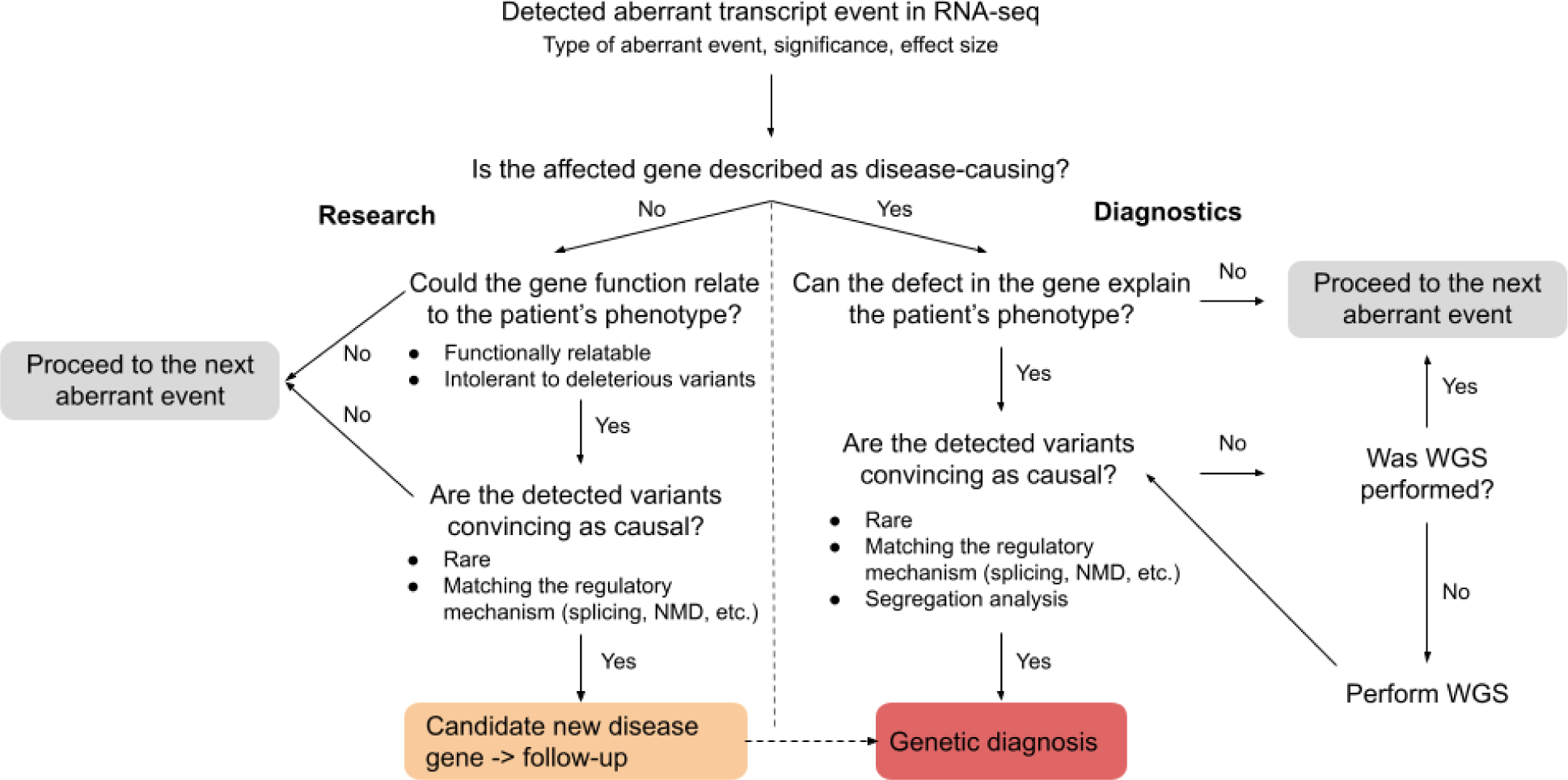
RNA-seq based diagnostic flow chart. Flow diagram showing the diagnostic decision guideline after detecting a gene with an aberrant event in RNA-seq data. Identification of an aberrant event can lead to genetic diagnosis (diagnostic setting), lead to discovery of a candidate new disease gene (research setting), or alternatively be of unlikely diagnostic significance, after which the next aberrant event is analyzed following the same way path.

### Aberrant Expression

A total of 14,100 genes were considered in the strand-specific subset and 14,399 in the non-strand-specific subset (fig. S3, coverage filter, Materials and Methods). In both cohorts, this represented 66% of the OMIM genes and 90% of the mitochondrial disease genes (Materials and Methods). OUTRIDER called a median of two underexpression outliers per sample, including one known disease gene, and a median of one overexpression outlier per sample, at a false discovery rate (FDR) less than 0.05 (Fig. 3A).

**Fig. 3.**
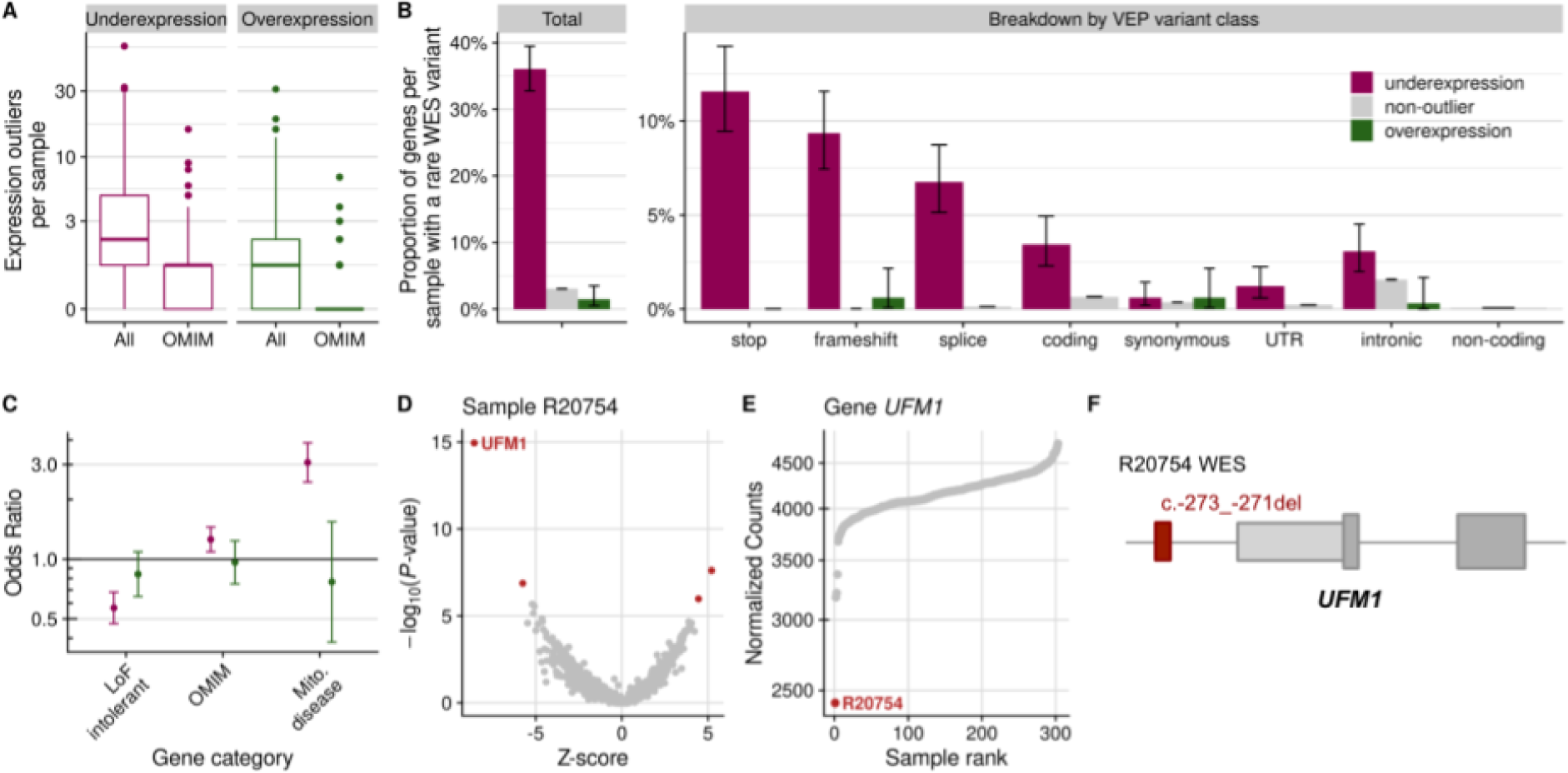
Aberrant expression. **(A)** Distribution of genes per sample that were detected as expression outliers, for all genes and genes known to cause a disease (OMIM), stratified by outlier class. **(B)** Proportion of genes per sample with a WES rare variant among underexpression outliers, non-outliers, and overexpression outliers, for all rare variants and stratified by VEP variant classes (Methods). Error bars represent 95% confidence intervals of a binomial test. **(C)** Observed over expected number of overexpression and underexpression outliers (y-axis, log-scale) for loss-of-function intolerant genes, OMIM genes, and mitochondrial disease genes (x-axis). Error bars represent 95% confidence intervals of pairwise logistic regressions. **(D)** Gene-level significance (-log_10_(*P*), y-axis) versus Z-score, with *UFM1* labeled among the expression outliers (red dots) of sample R20754. **(E)** Expression of *UFM1* shown as normalized counts ranked across all samples, with the lowest expression in sample R20754. **(F)** Schematic depiction of the NM_016617.2:c.-273_-271del *UFM1* deletion (red rectangle) detected by WES in sample R20754. Figure not shown at genomic scale. LoF: loss-of-function, VEP: variant effect predictor.

We systematically assessed how expression outliers are associated with classes of rare variants and classes of genes (Materials and Methods). We found at least one rare (minor allele frequency < 0.001) variant in 36% (293 out of 812) of underexpression outlier genes. This is a significantly higher proportion than what was observed in the overexpression and non-outlier genes, indicating a causal relationship (Fig. 3B). Notably, in 28% (225 out of 812) of the underexpression outliers, the rare variant could directly (stop, frameshift) or indirectly (splice region - within 1-3 bases of the exon or 3-8 bases of the intron) lead to a premature stop codon, triggering the nonsense-mediated decay (NMD, Fig. 3B). This is in agreement with previous reports describing the association between rare variants with aberrant expression in affected and non-affected individuals across various tissues (25,60–63). The remaining rare variants associated with the underexpression outliers were significantly enriched for coding, intronic, and variants in the untranslated regions (UTRs), indicating important regulatory roles of variants in these regions (Fig. 3B).

In agreement with observations in non-affected individuals (62), loss-of-function (LoF) intolerant genes were depleted for underexpression outliers (Fig. 3C), reflecting constrained expression. Given the clinical diagnosis of the individuals in our study, we observed an enrichment for OMIM genes (1.25-fold), and particularly mitochondrial disease genes (3-fold) among underexpression outliers (Fig. 3C). Overexpression outliers did not show an association with these gene categories, indicating them to be less clinically relevant. Collectively, these results support detection of underexpression outliers to guide towards a molecular diagnosis.

For candidate gene prioritization, we focused on underexpression rather than overexpression outliers because we presumed LoF to be a more likely pathomechanism of congenital metabolic disorders than dominant negative and gain-of-function (31). Aberrant expression was a major contributor to our diagnostic success with 25 out of 32 (78%) newly diagnosed cases pinpointed as expression outliers (fig. S4 and S5). In figure 3 D-F we illustrate how expression outlier detection supported the identification of a causative variant in the case of a male with neonatal-onset leukodystrophy, nystagmus, and hearing impairment, whose initial WES analysis was inconclusive. RNA-seq analysis revealed two underexpression outliers, among which was the ubiquitin-fold modifier 1 gene, *UFM1* (MIM: 610553, Fig. 3D). In this sample, we found the lowest expression of *UFM1* among all 303 samples (Fig. 3E). Reinspection of WES revealed a 3-bp homozygous deletion located in the promoter region (NM_016617.2:c.-273_-271del, Fig. 3F), which was initially not prioritized during WES analysis due to its location. This variant has recently been reported to significantly reduce promoter and transcriptional activity and was considered to be pathogenic in cases of hypomyelinating leukodystrophy (64). This case exemplifies how the detection of aberrant expression enables the reprioritization of variants located in the non-coding regions.

Detection of aberrant expression can directly pinpoint the causative gene, but it can also reflect downstream effects, which provides functional evidence and can guide or support diagnostic interpretation. This is exemplified in a case diagnosed after WES identified a 3-bp homozygous deletion (NM_133259.3:c.2595_2597del, p.Val866del) in the *LRPPRC* gene (MIM: 607544, (41)), encoding for a leucine-rich PPR motif-containing protein that forms a ribonucleoprotein complex with SLIRP to regulate the stability of mature mitochondrial transcripts (65). OUTRIDER detected a significant downregulation of 11 mitochondrial DNA (mtDNA)-encoded genes in this case (fig. S6), thus supporting the functional defect of LRPPRC, and contributing to the enrichment of mitochondrial disease genes among underexpression outliers.

### Aberrant Splicing

Aberrant splicing can be caused by variants in the canonical splice sites, but also by variants in weak splice sites or in less clearly mapped splicing regulatory sequences such as the exonic and intronic splicing enhancers and silencers (66). The aberrant splicing caller FRASER (56) is based on annotation-free intron-centric metrics (67). Specifically, FRASER uses percent-spliced-in of alternative donor sites (*Ψ*_5_) and alternative acceptor sites (*Ψ*_3_) to detect exon skipping, exon creation, exon truncation, and exon elongation, in addition to splicing efficiency (*θ*) to detect intron retention (Materials and Methods). After applying FRASER with the recommended cut-offs, we obtained a median of 23 genes with at least one aberrantly spliced junction per sample (FDR < 0.1 and differential *Ψ*_5_, *Ψ*_3_, or *θ* greater than 0.3), including 7 disease genes (Fig. 4A).

**Fig. 4.**
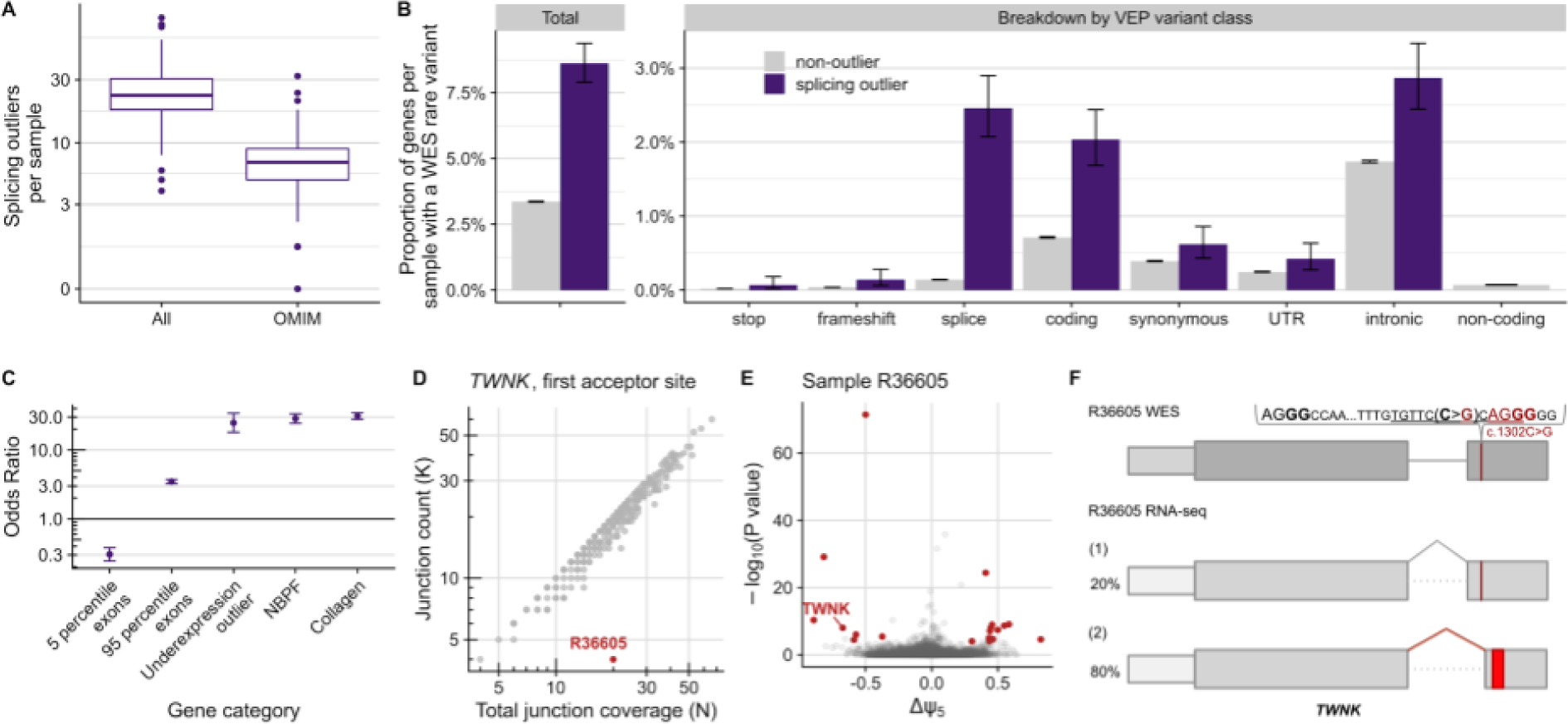
Aberrant splicing. **(A)** Distribution of genes per sample that had at least one splicing outlier, for all genes and genes known to cause a disease (OMIM). **(B)** Proportion of genes per sample with a WES rare variant among the splicing outliers (purple) and the non-outliers (gray), for all rare variants and stratified by variant effect predictor classes. Error bars represent 95% confidence intervals of a binomial test. **(C)** Observed over expected number of splicing outliers on different gene categories. NBPF and collagen genes were chosen due to their high number of exons and due to collagen genes alternative splicing in a developmental-stage manner (68) and NBPF genes having a repetitive structure, which exposes them to illegitimate recombination (69). Error bars represent 95% confidence intervals of pairwise logistic regressions. **(D)** Split-read counts (y-axis, gray junction on panel F) of the first annotated junction of *TWNK* against the total split-read coverage (x-axis, gray and red junctions on panel F) of the first donor site of *TWNK*. Many samples are not exclusively using the annotated junction (scattered below the diagonal), leading to a reference *Ψ*_5_ for the annotated junction of 87%. The observed *Ψ*_5_ for the first acceptor site of *TWNK* in the outlier sample is 20% (obtained by dividing the junction reads by the total junction coverage, 4/20). **(E)** Gene-level significance (-log_10_(*P*), y-axis) versus differential splicing effect (observed minus expected usage proportion of the tested donor site, Δ*Ψ*_5_, x-axis) for the alternative splice donor usage in sample R36605. Gene-level significance is obtained by multiple testing correction across all introns per gene (56). Outliers are marked in red and the gene *TWNK* explicitly labeled. The Δ*Ψ*_5_ value for the first donor site of *TWNK* in this sample is -0.67=0.2-0.87. **(F)** Schematic depiction of the synonymous NM_001163812.1:c.1302C>G (p.Ser434=) *TWNK* variant and its consequence on the RNA level, activating a new acceptor site (ACGG in red). and leading to the creation of a premature termination codon (red rectangle) in sample R36605. The percentage of each transcript isoform is shown next to it. Figure not shown at genomic scale.

In our cohort, 8.5% (487 out of 5,649) of aberrantly spliced genes contained rare variants, a proportion significantly higher than non-splicing outlier genes (Fig. 4B). As expected, many of these rare variants are located in the splice region (60,61). Moreover, we observed an enrichment of coding and intronic variants (Fig. 4B), underscoring the role of coding and intronic sequences in splicing. Although we controlled for multiple testing within genes (56), genes with more exons (n > 95^th^ percentile) had enrichment of aberrant splicing, while genes with fewer exons (n < 5^th^ percentile) showed less aberrant splicing (Fig. 4C). In particular, the neuroblastoma breakpoint family (NBPF) of genes, which have 35 exons per gene, and collagen genes, which have 54 exons per gene, compared to a median of 17 exons in genes detected by FRASER, appeared to be more susceptible to aberrant splicing. In addition, splicing outliers were found to be 25-fold enriched among underexpression outliers (Fig. 4C), some of which could be explained due to the creation of an aberrantly spliced isoform containing a premature termination codon (PTC) ultimately resulting in transcript degradation by NMD.

Aberrant splicing was considered disease-causative in 18 out of 32 cases (56%), 11 of which were in combination with supportive aberrant expression data (fig. S4C). In Fig. 4D to F, we showcase how aberrant splicing detection helped identify the disease-causing gene defect in a male patient with early-onset acute liver failure. In this sample, 38 aberrantly spliced genes were identified by FRASER, 13 of them on disease-causal genes. Among them was aberrant splicing and expression of *TWNK* (MIM: 606075). This gene encodes the twinkle mtDNA helicase that is critical for the efficient mtDNA replication and synthesis of the nascent D-loop strands (70). Autosomal recessive pathogenic variants in *TWNK* are associated with disorders of mtDNA maintenance, including a hepatocerebral presentation associated with a mtDNA depletion (71). FRASER called a significant deviation from the canonical junction usage of the first intron (Fig. 4D), whereby an alternative acceptor within exon 2 was utilised, resulting in a frameshift and creation of a premature termination codon (Fig. 4F). Moreover, this aberrant splicing was associated with reduced level of the *TWNK* transcript (fold change: 0.43) indicating NMD of some transcript isoforms. Reanalysis of WES data revealed a rare homozygous variant (NM_001163812.1:c.1302C>G) in the second exon, which is predicted to have a synonymous effect (p.Ser434=). The variant is positioned four nucleotides upstream of an alternative splice junction (Fig. 4F), which corresponds to a weak splice site in controls (*Ψ*_5_ = 9%). This variant is predicted (using the Human Splicing Finder *in silico* tool (72)) to disrupt the plausible exonic splicing enhancer sequences tgttcCca and ccCagg (Fig. 4F), thereby activating the weak splice site and eventually leading to a frameshift (p.416Glyfs*7). The activation of weak splice sites is a likely disease-causing phenomenon and is known to be recurrent, as we reported in an earlier study (23).

A particular value of RNA-seq lies in the quantification of different transcript isoforms. This is especially useful for transcripts with physiological presence of several alternative isoforms and in cases of aberrant splicing with a complex pattern, exemplified by a case with a homozygous splice region variant (NM_022915.3:c.179+3A>G) in the gene *MRPL44* (MIM: 611849). This variant led to transcript depletion and three alternative isoforms with a PTC in each, in addition to the main transcript isoform that was present in less than 18% of all reads (fig. S7).

### Mono-allelic expression

For heterozygous loci, the expression of only one of the two alleles is referred to as mono-allelic expression (MAE). Possible causes for MAE include one allele being transcriptionally silenced or post-transcriptionally degraded and can have genetic or epigenetic grounds (73,74). As heterozygous variants alone are discarded when investigating autosomal recessive disorders, the detection of MAE of a rare variant indicates that both alleles are affected and enables their prioritization. We call MAE on heterozygous single nucleotide variants (SNVs) with at least 10 counts (median: 8,542 per sample, Fig. 5A) using the negative binomial test of Kremer et *al.* (23) (FDR<0.05 and allelic imbalance >80%, Material and Methods). We observed that MAE was more frequent towards the reference (median of 434) than towards the alternative allele (median 118, Fig. 5A). Subsetting to rare variants we found a median of 30 mono-allelic events towards the reference allele and 1 towards the alternative (Fig. 5A).

**Fig. 5.**
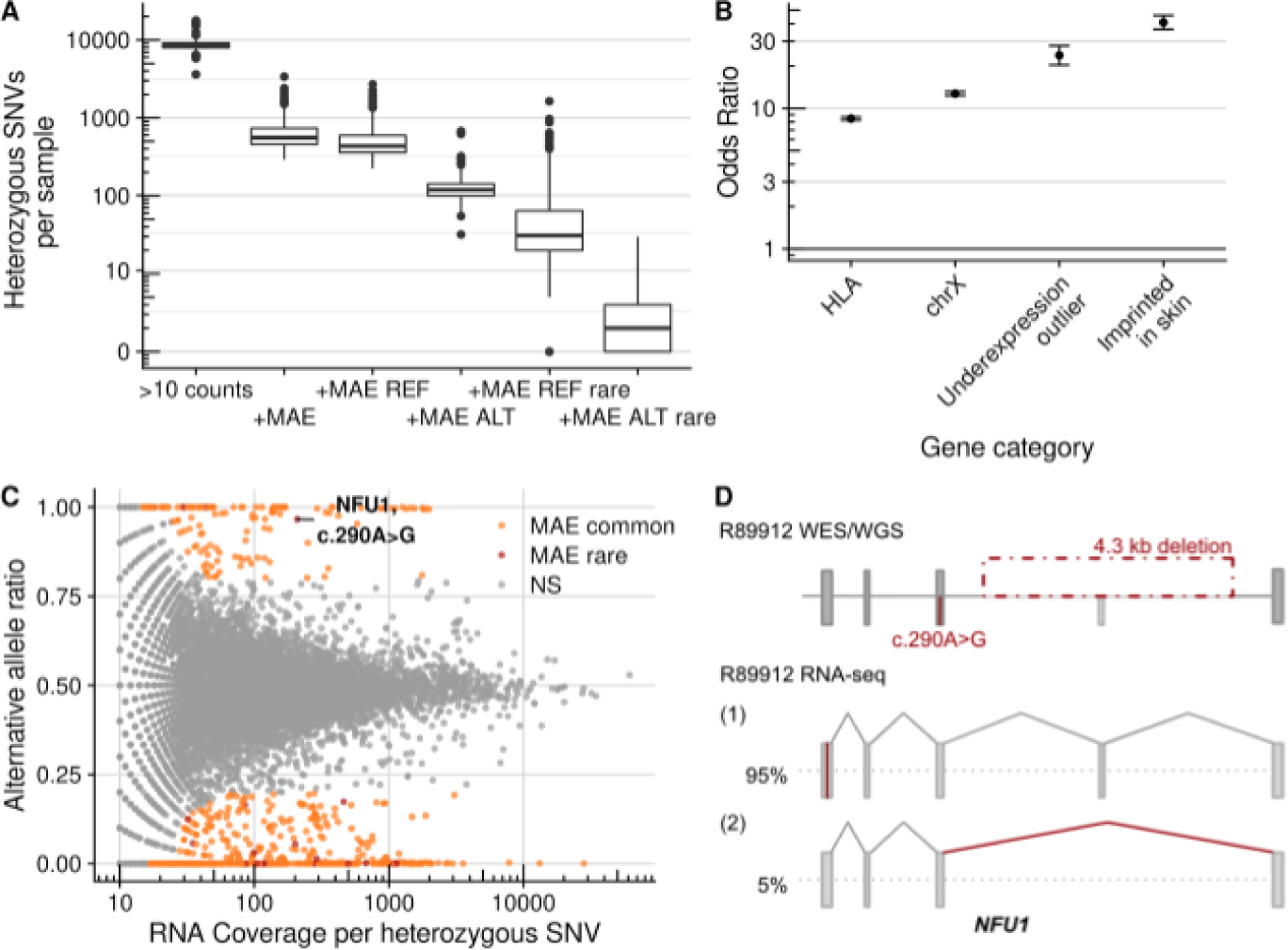
Mono-allelic expression. **(A)** Distribution of heterozygous SNVs per sample for successive filtering steps from left to right: Heterozygous SNVs detected by WES with an RNA-seq coverage of at least 10 reads, where MAE is detected, where MAE of the reference is detected, where MAE of the alternative is detected, and subsetted for rare variants. **(B)** Odds ratio of MAE in genes with common variants only and with at least one rare variant across different gene categories. Error bars represent 95% confidence intervals of pairwise logistic regressions. **(C)** Alternative allele ratio (y-axis, alternative allele counts / total allelic counts) for the sample R89912 compared to the total allelic counts per SNV (x-axis) within the sample. Significant MAE in common variants are shown in orange and in rare variants in red, among which is the disease-causing SNV NM_001002755.2:c.290A>G *NFU1*. **(D)** Schematic depiction of the disease-causing 4.3 kb deletion and the c.290A>G SNV in *NFU1* and their consequence on the RNA level in sample R89912. The percentage of detected transcript isoform is shown next to it. Figure not shown at genomic scale. REF: reference, ALT: alternative, rare: minor allele frequency < 0.1%, NS: not significant.

As expected from their biology, MAE was enriched among HLA, X-chromosomal, and imprinted genes (Fig. 5B). Moreover, we found a high enrichment of underexpression outliers (Fig. 5B), indicating that loss of expression of a single allele can be detected as an aberrant expression of the gene itself.

Detection of MAE helped to diagnose four cases, all coupled with aberrant expression (fig. S4). In one case, an infant male with severe Leigh syndrome and complex I deficiency, MAE of *NFU1* (MIM: 608100) was identified (Fig. 5C) in combination with aberrant expression (fold change: 0.63, fig. S5). *NFU1* encodes a scaffold protein that facilitates the insertion of iron-sulfur clusters into the subunits of the respiratory chain complexes and lipoic acid synthase (75,76). Individuals harboring pathogenic biallelic *NFU1* variants present with early-onset failure to thrive, pulmonary hypertension, encephalopathy, and neurological regression (77,78). The mono-allelically expressed missense variant NM_001002755.2:c.290A>G (p.Val91Ala) with a CADD score of 28.6 was absent from gnomAD. Following this observation we initiated WGS, which revealed a 4.3 kbp heterozygous deletion affecting exon 6 of the second allele, explaining the detected MAE (Fig. 5D). Proteomic analysis also found a severe reduction of NFU1 (fold change: 0.13) finally confirming the diagnosis (59). This case presents the first association of pathogenic *NFU1* variants with Leigh syndrome, thus expanding the clinical phenotype. Interestingly, pathogenic variants in another iron-sulfur cluster scaffold gene, *BOLA3* (MIM: 613183), and in a gene involved in iron-sulfur cluster biosynthesis, *FDXR* (MIM: 103270), have previously been reported to cause Leigh syndrome (79,80).

Haploinsufficiency has been reported as a pathomechanism for more than 660 genes (81,82). It appears especially important for neurodevelopmental disorders, where *de novo* variants are often found in haploinsufficient genes or regulatory non-coding regions (83,84). Hence, although the majority of mitochondrial diseases are inherited in an autosomal recessive mode (31), we also considered the possibility of haploinsufficiency, which can be detected by a combination of underexpression and MAE. Three samples in our cohort were solved with known haploinsufficient genes (*MEPCE* (MIM: 611478, ref. (45)), *SON* (MIM: 182465), and *CHD1* (MIM: 602118), Table S3). They were all called outliers with close to 50% reduction in expression level (fold change: 0.56, 0.61, and 0.64, respectively, fig. S5). Each of these genes had heterozygous protein-truncating variants suggesting that NMD acted on the transcript originating from the alternative allele. This hypothesis was confirmed by MAE of the reference alleles (88%, 91%, 75%, respectively). Moreover, segregation analysis in each case confirmed that the protein-truncating variants occurred *de novo*. Altogether, these results demonstrate that aberrant expression callers controlling for hidden confounders such as OUTRIDER (55) are sufficiently sensitive to detect aberrant expression when only one allele is affected and may discover the pathological variant in autosomal dominant disorders, particularly those presenting haploinsufficiency.

### Variant calling in RNA-seq data

Currently, the application of WGS is still emerging within a diagnostic setting, largely limiting the sequence analysis to coding variants detected by WES. Advantageously, RNA-seq is able to contribute to variant discovery (26) by covering UTRs and even, with a lesser sequencing depth, intronic regions, which are not well covered by exome-capturing kits (85). We called variants in our RNA-Seq data following GATK’s best practices (Materials and Methods). To identify filtering criteria with a useful balance between recall and precision, we performed a benchmark using 210 RNA-Seq samples derived from suprapubic skin of the GTEx project (86), considering their corresponding WGS-based variants as the ground truth. This benchmark suggested the exclusion of regions with three or more variants within a 35 bp window, repeat masked regions (87), and variants with less than three reads supporting the alternative allele. Filtering these variants yielded a precision of 95% (97%) and a recall of 40% (54%) for heterozygous (homozygous) variants, in genomic positions with an RNA coverage of at least three reads (fig. S8A, Materials and Methods). While a precision of 95% would not be recommended for genome-wide variant prioritization, we found it to be reasonable for variant detection in candidate genes identified by aberrant expression and splicing analyses. When applied to our rare disease compendium, these filters yielded a median of 44,154 variants per RNA-seq sample, in comparison to a median of 63,666 variants found by WES (fig. S8B), including a median of 19,252 variants not called by WES. As expected, RNA-seq was particularly helpful in revealing variants in the untranslated regions (40% RNA-seq only in 5’UTR and 75% in 3’UTR, Fig. 6A). Coverage of intronic regions increased by one third by using RNA-seq based calling, which was specifically helpful for detecting deep intronic splice-altering variants (Fig. 6B).

**Fig. 6:**
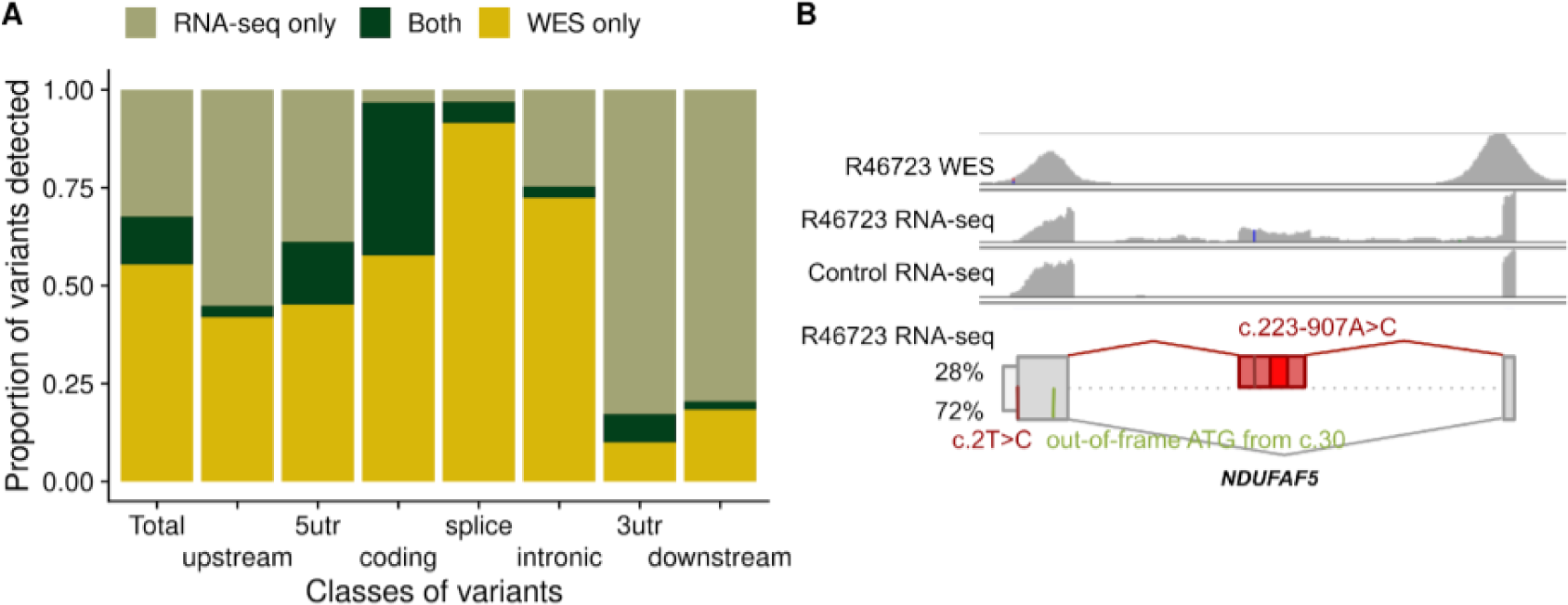
RNA-seq variant calling. **(A)** Median across samples of the proportion of variants called only by WES, only by RNA-seq, and by both technologies, in total and stratified by variant classes. Of note, over 50% of variants in coding regions are called only by WES, probably because of RNA-seq limitations including that not all the genes are expressed in fibroblasts, the uneven read coverage along the transcript, and because the expression level of variant-carrying alleles must be high enough to yield sufficient RNA-seq read coverage. **(B)** Rows 1 - 3: WES and RNA-seq coverage of the affected sample (R46723) and a representative control using IGV. The created exon and a variant are seen in the affected RNA profile, but not covered in the corresponding WES and not present in the control. Bottom row: schematic depiction of the NM_024120.4 c.2T>C and c.223-907A>C variants and their consequence on the RNA level with an out-of-frame ATG (in green), and a cryptic exon with the PTC (bright red rectangle) on *NDUFAF5*. The percentage of detected transcript isoform is shown next to it. Figure not shown at genomic scale.

RNA-seq variant calling identified the causative variant in nine cases missed by WES, all deep intronic (Table 1 and Fig. 6B), and thereby negating the need to perform WGS. All of these cases were already candidates identified via aberrant expression and/or splicing analyses. One was a female suspected mitochondrial disease patient, presenting early in infancy with failure to thrive, complex I deficiency, and elevated lactate. FRASER detected aberrant splicing in the *NDUFAF5* gene (MIM: 612360), encoding a complex I assembly factor, highlighting a cryptic exon in intron 1, present in 28% of the transcript (Fig. 6B). This 258-nt cryptic exon is in frame and predicted to lead to an extension of the open reading frame by 31 amino acids before encountering a PTC (Fig. 6B). RNA-seq variant calling revealed a rare intronic variant (NM_024120.4:c.223-907A>C) within the cryptic exon (Fig. 6B). This variant has recently been described in a single patient, with cDNA studies supporting the creation of a new exonic splicing enhancer and the same aberrant splicing (88). Moreover, WES had identified an unreported start-loss heterozygous variant (NM_024120.4:c.2T>C). This variant disrupts the start codon, with the next available ATG out-of-frame at position c.30. Pathogenic variants in *NDUFAF5* have been associated with an early-onset complex I deficiency, characterized by developmental delay, failure to thrive, hypotonia, and seizures (88), in agreement with the clinical presentation of the investigated individual. This intronic variant was also found associated with the inclusion of the same cryptic exon in another unrelated RNA-seq diagnosed case from our compendium, where it is *in trans* with a heterozygous frameshift NM_024120.4:c.605dup which causes aberrant expression (Table 1). Notably, variant calling in RNA-seq data fails in intergenic and intronic regions, as well as in genes that are not expressed. Thus, the increased power of WGS in calling all genetic variation is still unquestionable, though the interpretation of cumbersome WGS datasets could be streamlined through the incorporation of RNA-seq data (Fig. 2).

### Overview of the diagnosed cases

In a diagnostic setting, the value of RNA-seq lies in the functional assessment of often unpredictable effects of variants, leading to their validation and (re)prioritization, or shedding light on the non-coding regions and more complex pathomechanisms. As seen from the 32 cases diagnosed using RNA-seq, this application enabled the detection of a broad spectrum of molecular pathomechanisms driven by rare variants, including aberrant expression caused by variants in the promoter, deep intronic variants generating cryptic exons, and the combined deleterious effect of two common variants in *cis* (Table 1, Fig. 7).

**Fig 7.**
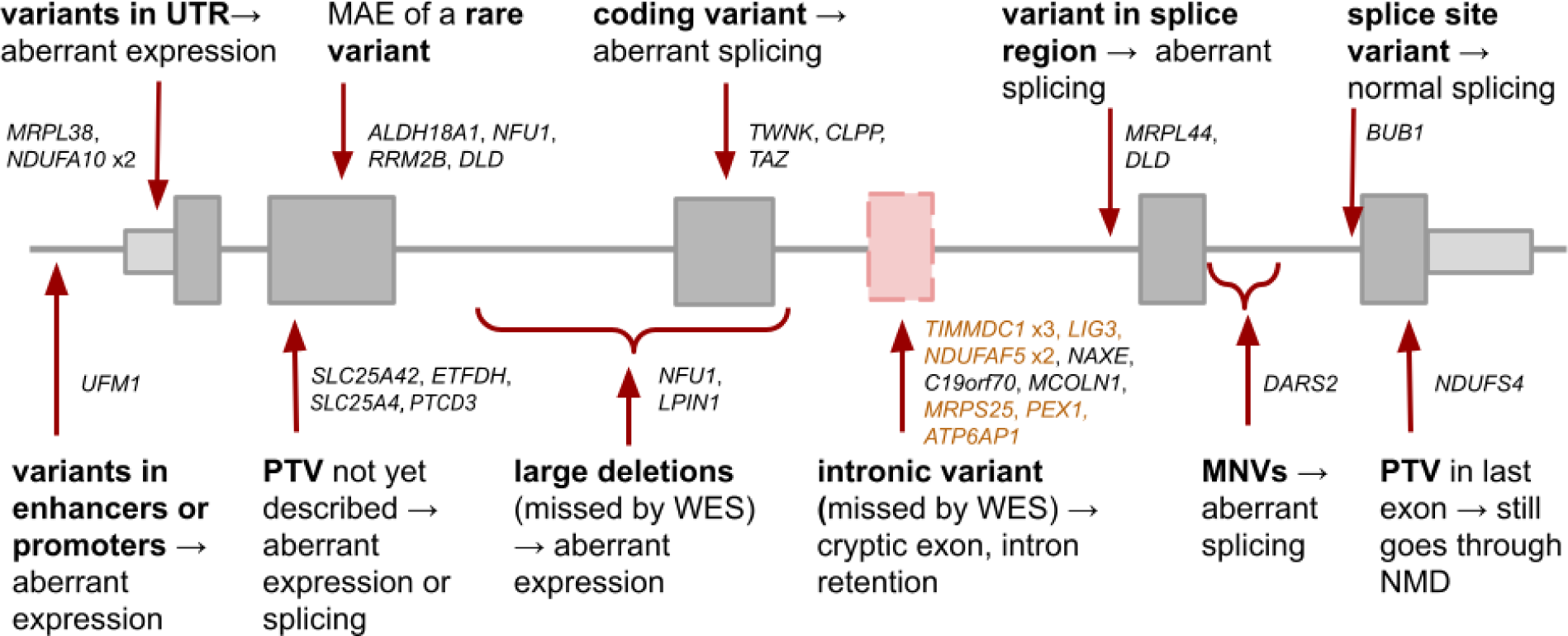
RNA-seq captures a broad spectrum of mechanisms of action of pathogenic variants. Summary of variants and their effect on transcript across 33 cases from our cohort, where the capture of a transcript event by RNA-seq enabled establishing genetic diagnosis in 32 and rejecting a candidate gene in one case, highlighting the value of transcriptomics as a tool in diagnostics. Each gene represents one case, except for *NFU1* that belongs to two categories. Highlighted in orange are the nine cases where the intronic variant was missed by WES, but called by RNA-seq. Both large deletions were missed by WES and RNA-seq, therefore requiring WGS to be identified. PTV: protein-truncating variant. MNV: multi-nucleotide variant.

Returning to our reference set, of the 98 WES-diagnosed cases, 31 contained at least one rare protein-truncating variant (PTV) (fig. S1, table S3). Our RNA-seq-based workflow was able to re identify the causal gene in 84% (26 out of 31) of them (fig. S1, table S3). In the five remaining cases, RNA-seq failed to detect the aberrant transcripts for various reasons. In two cases, the causal genes (*CCDC40*, MIM: 613799; and *F11*, MIM: 264900) were not expressed in skin fibroblasts. In another two cases, both involving the *TXNIP* gene (MIM: 606599), the fold change was low (0.1 and 0.22), but a high dispersion of the gene’s expression led to the cases not being called as outliers. In the last case, the patient harbored a heterozygous pathogenic missense variant *in trans* with a heterozygous PTV located in the first exon of the *GFER* gene (MIM: 600924) that leads to a frameshift and PTC after 16 amino acids. The clinical presentation of the patient and his sibling (not included in our cohort) include exercise intolerance, elevated lactate, and cataracts, which matched those described in other cases harbouring biallelic *GFER* variants (89), arguing in favor of pathogenicity. This case exemplifies that the functional impact of PTC is not always explained by NMD (90).

We inspected all rare homozygous variants and found that most of the rare stop and frameshift variants that occur outside the last exon and expressed in fibroblasts (Materials and Methods) indeed caused an expression outlier (fig. S9A). Only one rare homozygous frameshift in the last exon led to aberrant expression, which corresponded to the *NDUFS4* case (Fig. 7, fig. S5), thus providing functional validation to solve the case. Similarly, most homozygous splice-site variants caused aberrant splicing (fig. S9B), with the exception of one case where normal splicing was observed (variant NM_004336.4:c.1876+2A>G in *BUB1*, Fig. 7, fig. S9C). In this respect, RNA-seq can also help to prevent a false assignment of pathogenicity. Finally, we tested the impact of all rare stop heterozygous SNVs that are not located in the last exon and found that more than 70% led to MAE of the reference allele (fig. S9D), which was a significantly higher proportion than the rest of variant classes and is in agreement with the results obtained from GTEx (61,91). This highlights the need for functional validation, such as RNA-seq, for PTV without reported pathogenicity (Fig. 6).

Reanalysis of the cohort published in 2017 (23) provided a genetic diagnosis to an additional five cases (two *NDUFAF5*, *LPIN1*, *TAZ, NDUFA10*; Table 1). The splicing defect in *NDUFAF5* was not detected by the previously used LeafCutter method (92) but was found with FRASER. LeafCutter did find aberrant splicing in the *LPIN1* and *TAZ* genes, but no variants were found to be conclusive. For the *LPIN1* case, the causative variant (a 1,759 bp deletion) was detected in a follow-up study via WGS (Fig. 7). For the *TAZ* case, the causative variant was initially not prioritized because of its predicted consequence (56). In the *NDUFA10* case, the previously used DESeq2 (93) for aberrant expression did find it to be an expression outlier, but the homozygous causal variants in the 5’UTR were not initially prioritized until later when the same aberrant expression was identified in the affected sibling (fig. S5). This shows the importance of data reanalysis by considering updates of the disease course and family segregation, applying state-of-the-art methods, and follow-up studies.

### Tissue-specific gene expression

An important limitation of the application of RNA-seq in a clinical setting is that the causal gene may not be expressed in the sampled tissue. To assess the impact of source material on transcriptome, we compared the expression of disease-associated genes for major disease categories (Fig. 8, Materials and Methods) across 49 tissues from healthy donors from the GTEx Consortium (86). The majority of genes of each disease category are expressed in any given tissue (except for ophthalmology and skeletal dysplasia genes in whole blood, Fig. 8A). Exceptionally, mitochondrial disease genes are ubiquitously expressed in all tissues, but other disease genes have a more pronounced tissue-specific expression profile, such as neurological genes in the brain (Fig. 8A). As in clinical practice biopsy of the least invasive tissue is desirable, we next focused on the clinically accessible tissues (CATs) - whole blood, Epstein-Barr virus (EBV)-transformed lymphocytes, skeletal muscle, and skin-derived fibroblasts (94). Fibroblasts were the CAT expressing the highest number of Mendelian disease genes (2,564; 67%; Fig. 8B). Although obtaining a skin biopsy is more invasive, skin-derived fibroblasts appear as a more useful resource than blood, showing a higher number of expressed genes in each disease category (which is significant for OMIM, neurology, ophthalmology, and skeletal dysplasia, Fisher’s test *P*<0.05). Fewer genes are also expressed in muscle for all disorders, except for neurology and neuromuscular disorders, confirming its utility (Fig. 8B).

**Fig 8.**
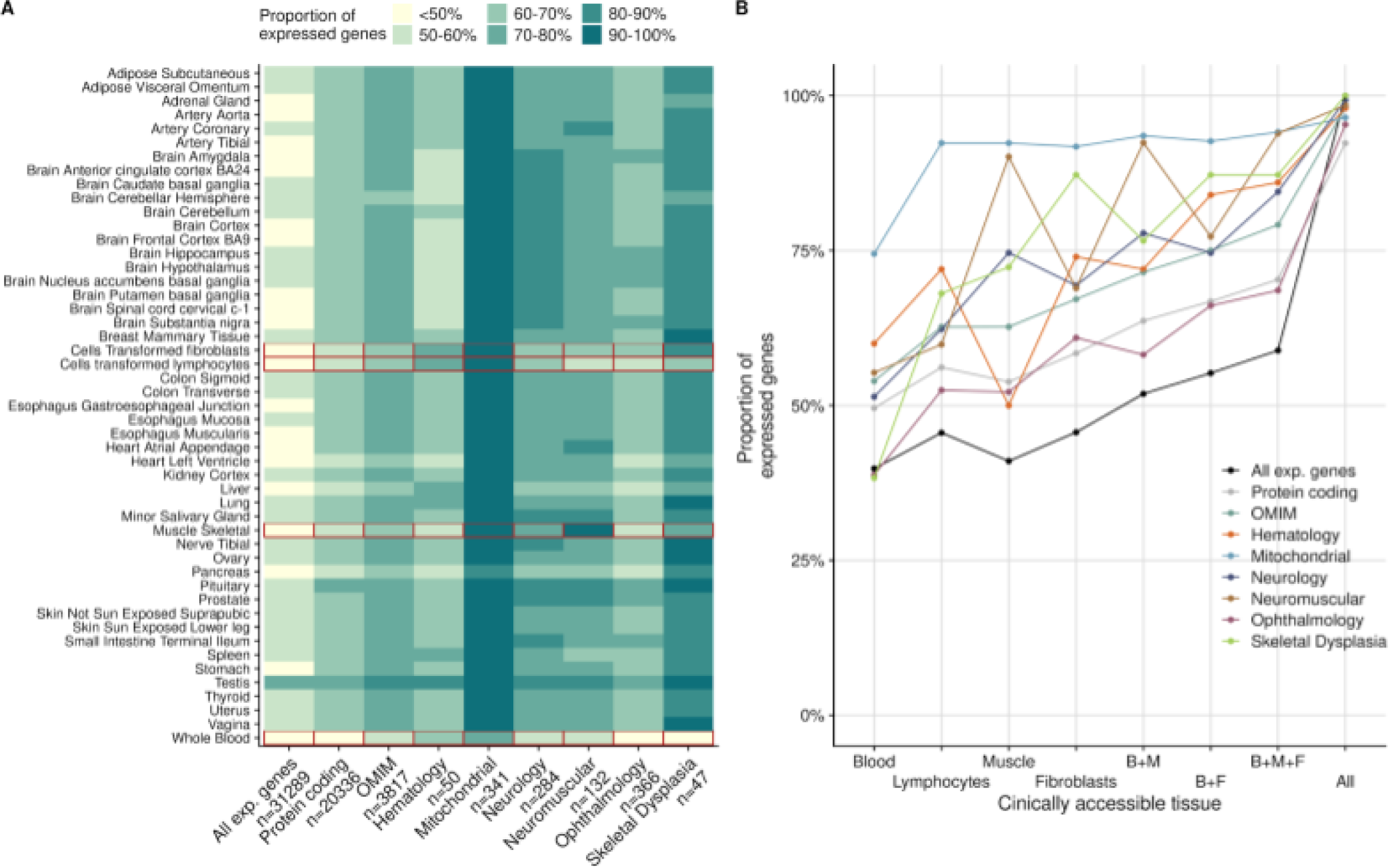
Tissue-specific gene expression. **(A)** Proportion of expressed genes from different categories across 49 GTEx tissues with the CATs delineated in red. **(B)** Proportion of expressed genes from different categories across CATs from GTEx, alone or in combination with another CAT. ‘All’ refers to all the 49 tissues, not just the CATs. B: blood, M: muscle: F: fibroblasts, CAT: clinically accessible tissue.

## Discussion

We report the outcome of RNA-seq implementation as part of routine diagnostics for Mendelian diseases alongside WES in our centre for more than 300 individuals. We demonstrate application of the automated computational workflow DROP, showcase detailed diagnostics successes including instances of dominant mode of inheritance, and provide a diagnostic decision workflow integrating WES, WGS, and RNA-seq. The computational analysis time for RNA-seq is comparable to the genome pipelines, typically requiring less than a week from sample preparation to reported results. RNA-seq is based on the same technology as WES/WGS, which is another favorable feature to consider when deciding to expand the diagnostic spectrum beyond the DNA sequence. Stringent p-value driven results yield a manageable number of OMIM genes with aberrant RNA events (median = 8), similar to the average number of biallelic rare non-synonymous variants inspected during diagnostic WES analysis of autosomal recessive disorders. In this study, cumulative evidence from WES and RNA-seq supported the genetic diagnosis in 16% of WES-inconclusive cases. This number falls within the range of other RNA-seq studies with unrestricted inclusion criteria ranging from 7.5-18% (fig. S10), and a hypothetical yield of 13.5% after retrospectively analyzing a cohort of WES-diagnosed patients (95), thereby reflecting the likely expected additional value of RNA-seq as a complement to WES.

The clinical significance of any given genetic variant falls along a gradient, ranging from those in which the variant is almost certainly pathogenic for a disorder, to those that are almost certainly benign (21). The clinical interpretation of variants is based on cumulative evidence from population-wide frequency, computational prediction, functional data, and segregation patterns. Findings from RNA-seq contribute evidence and may validate predicted pathogenic or likely pathogenic variants, reclassify VUS or benign variants, and lead to the detection of variants undetected by WES or overlooked by WES data inspection. Specifically, the value of RNA-seq is to provide functional evidence on variants affecting gene expression and splicing. Loss-of-function variation in disease genes represents the strongest evidence for pathogenicity. RNA-seq allows the detection of non-coding loss-of-function variation (96), including splice defects leading to the non-functional mRNA isoforms and variants abolishing transcription, which remain a challenge to predict from DNA sequence alone. Moreover, RNA-seq allows the validation or invalidation of (computational) predictions including splice site dinucleotide and NMD by providing quantitative measures of the actual fraction of affected transcripts. In this respect, RNA-seq could become the first step in the systematic inclusion of functional data together with genetic and phenotypic information which can relatively easily be implemented in the diagnostic workflow.

RNA-seq relies on taking additional patient biopsies in addition to a sample for DNA extraction and requires early consideration in the diagnostic process. Specifically, for severe life-threatening diseases with fast progression, we recommend establishing skin biopsies in the routine process in parallel to genome-based diagnostics. As we demonstrate, fibroblast cell lines express the majority of OMIM disease genes. Moreover, fibroblast cell lines can be differentiated into other cell types to more closely reflect the disease-affected tissue (26) or into pluripotent stem cells, where as many as 27,046 genes are expressed (63,97). Importantly, patient-derived cell lines not only allow functional studies of the disease of the patient but also provide a DNA resource for emerging sequencing technologies such as long read sequencing (98). One limitation of cell lines, in contrast to direct biopsies such as whole blood samples, is the time and effort needed for their growth. Therefore, in urgent situations (e.g., neonatal cases), blood sampling is preferred as it can be processed immediately.

There have been concerns regarding the use of fibroblasts to study pathological effects in diseases not affecting the skin. One concern is that the gene or its relevant isoform may be not expressed in fibroblast cell lines. Resources such as GTEx, Panel Analysis of Gene Expression (PAGE, (26)), or MAJIQ-CAT (94) allow expression of candidate genes and isoforms to be checked in clinically accessible tissues and cell types. Here, using GTEx, we demonstrate skin-derived fibroblasts to capture the majority of disease genes for major disease categories. Moreover, so long as the potential causal genes are expressed, non-affected tissues have the advantage that the transcriptome-wide consequences of the diseases are limited and hence the causal defects can more clearly stand out. Another concern is that pathogenic variants affect tissue-specific regulatory elements such as transcription factor binding sites or binding sites of tissue-specific splicing factors. However, strong regulatory effects, which one could expect for monogenic disorders, may rather be constitutive. This is the case for NMD (91). Also, a minor proportion of variants found in GTEx associated with splicing show tissue-specific effects (99). Overall, more studies are needed to assess the proportion of pathogenic variants altering tissue-specific regulatory elements.

With increasing adoption of RNA-seq in Mendelian disease diagnostics, we foresee the need for extended clinical guidelines, akin to the update of the ACMG/AMP guidelines necessitated by the uptake of WES as a standard diagnostic approach (21). These extended guidelines will need a concerted discussion across the community, regarding effect sizes and statistical cutoffs to define a pathological expression phenotype. Moreover, community-accepted criteria will be needed to assign the likelihood of pathogenicity for genomic variants, integrating RNA-seq based evidence with current features including annotations of genetic variants, computational predictions, frequency, and segregation patterns.

## Materials and Methods

### Compendium

A total of 303 individuals with a suspected mitochondrial disorder were recruited (Table S1). WES and RNA-seq were performed in all of them. The compendium includes 70 individuals from the Kremer et al. study (23), 152 individuals from a multi-omics study (59), and 81 additional individuals recruited for this study. Three cases further required WGS as they were candidates from RNA-seq, but no conclusive variants were found via WES or RNA-seq.

### Cell culture

Primary fibroblast cell lines obtained from patient skin biopsy were cultured in high glucose DMEM (Life Technologies) supplemented with 10% FBS, 1% penicillin/streptomycin, and 200 μM uridine at 37 °C and 5% CO_2_. All fibroblast cell lines tested negative for mycoplasma contamination.

### Whole exome sequencing

DNA was isolated from peripheral blood leukocytes or skin-derived fibroblasts using DNeasy Blood & Tissue Kit (Qiagen, Hilden, Germany) according to the manufacturer’s protocol. DNA concentration was measured using the Qubit™ dsDNA BR Assay Kit. 3 µg of DNA was used for library preparation. Exonic regions from human DNA samples were enriched with the SureSelect Human All Exon V5/V6 kits from Agilent (Agilent Technologies, Santa Clara, CA, USA) and sequenced as 100 bp paired-end runs on Illumina HiSeq2500 or HiSeq4000 platforms (Illumina, San Diego, CA, USA). Reads were aligned to the human reference genome (UCSC build hg19) using Burrows-Wheeler Aligner v0.7.5a (100). Single nucleotide variants, as well as small insertions and deletions (< 200 bp), were detected with SAMtools v0.1.19 (101) and GATK v3.8 (102).

### Whole genome sequencing

One WGS library was established using a MGIEasy DNA Library Prep Kit v1.1, according to the manufacturer’s protocol and generated DNA nanoballs. Sequencing was performed using 100-bp paired-end reads on a MGISEQ-2000 using MGISEQ-2000RS High-throughput Sequencing Set PE100 v3.0. The other two WGS libraries were prepared with the TruSeq DNA PCR-Free Kit (Illumina). DNA was fragmented to an average length of 350 bp by sonication. Libraries were validated according to standard procedures and sequenced via 150 bp paired-end on a NovaSeq 6000 platform. After removing adapter sequences and low-quality reads by Trimmomatic v0.39 (103), reads were aligned and variants were called using the same procedures described in the previous subsection.

### Variant annotation and handling

Variants were annotated for consequence, location, minor allele frequencies (from the 1000 Genomes Project (104) and gnomAD (105) cohorts), and deleteriousness scores using the R interface to the Ensembl Variant Effect Predictor (VEP) v1.32.0 (16,106). For variants that fell on multiple transcripts and had therefore multiple predicted consequences, the one with the highest predicted impact was selected (107). We considered a variant to be rare if the maximum minor allele frequency across both cohorts was lower than 0.001 and the frequency of the variant in our cohort was lower than 0.01. Variants are reported using the Human Genome Variation Society (HGVS) recommendations (108).

In order to detect whether a genomic position is expressed, we computed the RNA coverage using the coverage function from the GenomicAlignments R package v1.26.0 (109). We defined a position to be expressed if the mean coverage across all samples was greater or equal than 10 reads.

### RNA-sequencing

RNA was isolated from the patient-derived skin fibroblasts with the RNeasy mini kit (Qiagen, Hilden, Germany) according to the manufacturer’s protocol. RNA integrity number (RIN) was determined using the Agilent 2100 BioAnalyzer (RNA 6000 Nano Kit, Agilent Technologies, Santa Clara, CA, USA). Non-strand-specific RNA-seq was performed in 101 samples as previously described (23). The rest of the RNA samples were sequenced strand-specifically, where library preparation was performed according to the TruSeq Stranded mRNA Sample Prep LS Protocol (Illumina, San Diego, CA, USA). Specifically, 1 μg of RNA was purified using poly-T oligo-attached magnetic beads and fragmented. The RNA fragments were reverse transcribed with the First Strand Synthesis Act D mix. The second strand cDNA was generated with Second Strand Marking Mix that ensures strand specificity by replacing dTTP with dUTP. The resulting double-stranded cDNA was subjected to end repair, A-tailing, adaptor ligation, and library enrichment. The quality and quantity of the RNA libraries were assessed with the Agilent 2100 BioAnalyzer and the Quant-iT PicoGreen dsDNA Assay Kit (Life Technologies, Carlsbad, CA, USA). RNA libraries were sequenced as 100 bp paired-end runs on Illumina HiSeq2500 or HiSeq4000 platforms. Reads from RNA-seq were demultiplexed and then mapped with STAR (v2.7.0a) to the hg19 genome assembly, with default parameters plus setting the twopassMode to ‘Basic’ to detect novel splice junctions (110).

### Variant calling in RNA-seq data

Variants were called on RNA-seq data using GATK best practices for RNAseq short variant discovery (Online resources). Variants with a ratio of quality to depth of coverage < 2, that were strand biased (Phred-scaled fisher exact score >30), or belonging to a SNP cluster (3 or more SNPs within a 35 bp window) were filtered out, as suggested by GATK. Furthermore, variants not contained in a repeat masked region (as defined by RepeatMasker v4.1.0 (87)), and with 3 or more reads supporting the alternative allele were prioritized. For the benchmark analysis, 210 RNA-Seq samples derived from suprapubic skin of the GTEx project (86) were used. Only genomic positions with an RNA coverage of at least 3 reads were considered.

### Quality control

Quality control was performed using RNA-SeQC v2.4.2 (54) and visualized using MultiQC v1.9 1. (111) to detect the percentage of reads falling in exonic regions, and with low quality. DROP (v1.0.3) was used to compute the total sequencing depth per sample, percentage of mapped reads, and number of expressed genes (30). DROP was also used to determine whether an RNA-seq sample matches its annotated DNA sample. A cutoff of 0.7 distinctly separated the matching with the non-matching DNA-RNA pairs.

### Detection of aberrant expression

Detection of aberrant expression was fully based on DROP, v1.0.3 (30). We used as reference genome the GRCh37 primary assembly, release 29, of the GENCODE project (112) which contains 60,829 genes. We use the summarizeOverlaps function from the GenomicAlignments (109) R package to count reads that are paired with mates from the opposite strands (singleEnd = FALSE). We only considered reads that fell completely within an exon or span two exons from the same gene via splicing (mode = intersectionStrict). Reads that overlapped more than one feature were assigned to each of those features instead of being removed (inter.feature = FALSE). Genes with a 95th percentile FPKM < 1 were considered to be not sufficiently expressed and filtered out.

Expression outliers were found using OUTRIDER (54), a method that uses a denoising autoencoder to control for latent effects and returns multiple-testing corrected p-values (FDR) for each gene and sample. Significant events were defined as those with a FDR ≤ 0.05. All aberrant events were further inspected using the Integrative Genome Viewer (113).

### Detection of aberrant splicing

Splicing outliers were obtained using the DROP (v1.0.3) module based on FRASER (56), an annotation-free aberrant splicing detection algorithm. FRASER uses a denoising autoencoder to control for latent effects and estimates splice-site level and gene-level multiple-testing corrected p-values for percent-spliced-ins and splicing efficiencies. Exon-exon and exon-intron junctions with < 20 reads in all samples and for which the total number of reads at the donor and acceptor splice site is 0 in more than 95% of the samples were filtered out. From the FRASER output, splicing outlier genes were defined as those with a FDR ≤ 0.1 and an effect size (difference between the observed and the predicted percent-spliced-in, |Δψ| or between the observed and the predicted splicing efficiency, |Δθ|) larger than 0.3.

### Detection of mono-allelic expression

For mono-allelic expression analysis, only heterozygous single nucleotide variants from WES were considered. Reads assigned to each allele were counted using the ASEReadCounter function from GATK v4.0 (114). Positions with less than 10 reads in total were filtered out. Afterward, the negative binomial test described in Kremer *et al.* (23), was performed. This fully corresponds to the MAE module of DROP, v1.0.3.

### Association of outlier genes with WES rare variants

Variants were grouped by predicted consequence in a similar way as done in Li *et al.* (62), but with some minor modifications. Specifically, the categorization was: splice: splice acceptor, splice donor, splice region; frameshift: frameshift, UTR: 3’ UTR, 5’ UTR, start lost; non-coding: downstream, upstream, intron, regulatory region, intergenic; coding: coding, deletion, insertion, missense, stop lost; stop: stop gained; synonymous: synonymous, stop retained. Each sample-gene combination was categorized as overexpression, underexpression, or non-outlier according to the OUTRIDER results. Then, for each of them, we searched for a rare variant and assigned the variant’s consequence group to it. A Fisher’s exact test was performed for each variant group against each expression outlier class, thus obtaining a p-value. If rare variants from multiple groups were found on a sample-gene combination, the group with the lowest p-value (therefore highest association) was selected. Afterward, for each outlier class, the proportion of each group of rare variants was computed (e.g., # of underexpression outliers with a rare stop variant / total # of underexpression outliers). 95% confidence intervals were obtained from a binomial test for all proportions. The procedure was repeated in a similar way for splicing outliers caused by aberrant percent-spliced-in. Only protein-coding genes were considered. Samples with more than 20 expression outlier genes were discarded for the expression analysis, and samples with more than 40 splicing outlier genes were discarded for the splicing analysis.

### Association of WES rare variants with outlier genes

All rare variants in expressed protein-coding genes in autosomal chromosomes were considered. For each sample, each rare homozygous variant was matched with the corresponding outlier class (overexpression, underexpression, or non-outlier) of the gene where it is located. Then, for each group of rare variants, the proportion of each outlier class was computed (e.g., # of rare stop variants in a gene that is an underexpression outlier / total # of rare stop variants). Stop and frameshift variants that were in expressed positions and not in the last exon were marked as potential PTVs. The procedure was repeated in a similar way by associating rare variants with splicing outliers, but splitting the ‘splice’ category into ‘splice site’ (which includes both donor and acceptor dinucleotides) and ‘splice region’.

MAE was tested on each rare heterozygous SNV in genes in autosomal chromosomes. Then, for each group of rare variants, the proportion of each MAE category (towards the reference or alternative allele, or none) was computed (e.g., # of rare stop SNVs that are mono-allelically expressed towards the alternative allele / total # of rare stop SNVs).

### Enrichment of gene classes

We performed pairwise logistic regression where the response variable is the outlier class and the predictor the gene category. The odds ratio and 95% confidence interval were derived from the estimates and standard errors of the coefficients.

### Lists of genes

OMIM genes were downloaded from their portal (Online resources). Mitochondrial disease genes are our own expansion from the list shared in ref (31). Hematology, neurology, and ophthalmology genes were extracted from ref. (25), neuromuscular genes were taken from ref. (26), and skeletal dysplasia genes from ref. (28). Imprinted genes were taken from ref. (115). LoF intolerant genes correspond to the genes with a loss-of-function observed/expected upper bound fraction < 0.35 from ref. (105).

### Online resources

Code to reproduce the figures: https://github.com/gagneurlab/RNA_diagnostics_paper_figures

Pipeline to align and call WES/WGS variants: https://github.com/mri-ihg/ngs_pipeline/

DROP, https://github.com/gagneurlab/drop

GATK best practices for RNAseq short variant discovery, https://gatk.broadinstitute.org/hc/en-us/articles/360035531192-RNAseq-short-variant-discovery-SNPs-Indels-

GTEx Portal, https://www.gtexportal.org/home/

MAJIQ, https://majiq.biociphers.org/

OMIM, www.omim.org

PAGE, https://page.ccm.sickkids.ca

## Supplementary Material

fig. S1. Overview of the study. fig. S2. Quality control.

fig. S3. DNA-RNA sample matching. fig. S4. Aberrant events per sample. fig. S5. Expression fold change.

fig. S6. Case with many mtDNA expression outliers. fig. S7. Complex pattern of aberrant splicing.

fig. S8. Analysis on variants called by RNA-seq. fig. S9. Rare variants leading to outliers.

fig. S10. Diagnostic rate across cohorts.

table S1: Sample annotation.

table S2: Summary of candidate genes pinpointed via RNA-seq

table S3. Summary of WES-diagnosed cases with an RNA-defect

## Declarations

### Ethics approval and consent to participate

All individuals included or their legal guardians provided written informed consent before evaluation, in agreement with the Declaration of Helsinki and approved by the ethical committee of the Technical University of Munich.

### Consent for publication

Not applicable

### Availability of data and materials

The datasets (privacy-preserving count matrices) supporting the conclusions of this article are available without restriction in the Zenodo repository, doi: 10.5281/zenodo.4646823 and 10.5281/zenodo.4646827. The necessary data and the code to reproduce the main figures are available in GitHub: https://github.com/gagneurlab/RNA_diagnostics_paper_figures. Due to data privacy policy, we cannot provide BAM or VCF files.

## Authors’ contributions

Conceptualization: JG, HP. Data Curation Management: VAY, MG, RK, AN. Formal Analysis: VAY, MG, RK, CM. Investigation: MG, RK, AN. Resources: CLA, SB, HB, EC, FD, FF, PF, DG, JH, SJH, MH, YSI, YK, TK, TDK, CL, DL, CCM, JAM, SM, GMP, KM, AO, YO, DPA, EP, ARi, ARo, JS, CS, RWT, CT, FT, RVC, AV, MW, SBW, MX. Software: VAY, CM, NHS, MFM, RB, TS. Supervision: JG, HP. Validation: MG, RK, SLS, HP. Visualization: VAY, MG, JG, HP. Writing - Original Draft Preparation: VAY, MG, JG, HP. Writing - Review & Editing: all authors.

## Data Availability

Privacy-preserving count matrices are publicly available without restriction through Zenodo: 10.5281/zenodo.4646823 and 10.5281/zenodo.4646827. Due to data privacy policy, we cannot provide BAM or VCF files.

https://zenodo.org/record/4646823

https://zenodo.org/record/4646827

## Acknowledgements

We are grateful to the patients, their families, and the referring clinicians for participation in the study. We thank the Sequencing Core Facility of the HelmholtzZentrum München for providing the sequencing service. Figure 1 was created with BioRender.com.

## Abbreviations

CAT: Clinically accessible tissue
EBV: Epstein-Barr virus
FDR: False discovery rate
LoF: Loss-of-function
MAE: Mono-allelic expression
MNV: Multi-nucleotide variant
mtDNA: Mitochondrial DNA
NBPF: Neuroblastoma breakpoint family
NGS: Next generation sequencing
NMD: Nonsense-mediated decay
PTC: Premature termination codon
PTV: Protein-truncating variant
RNA-seq: RNA sequencing
SNV: Single nucleotide variant
UTR: Untranslated region
VEP: Variant effect predictor
VUS: Variant of uncertain significance
WES: Whole exome sequencing
WGS: Whole genome sequencing

