## Supplementary material for "Clinical implementation of RNA sequencing for Mendelian disease diagnostics": Sup Figures

### Supplementary figures

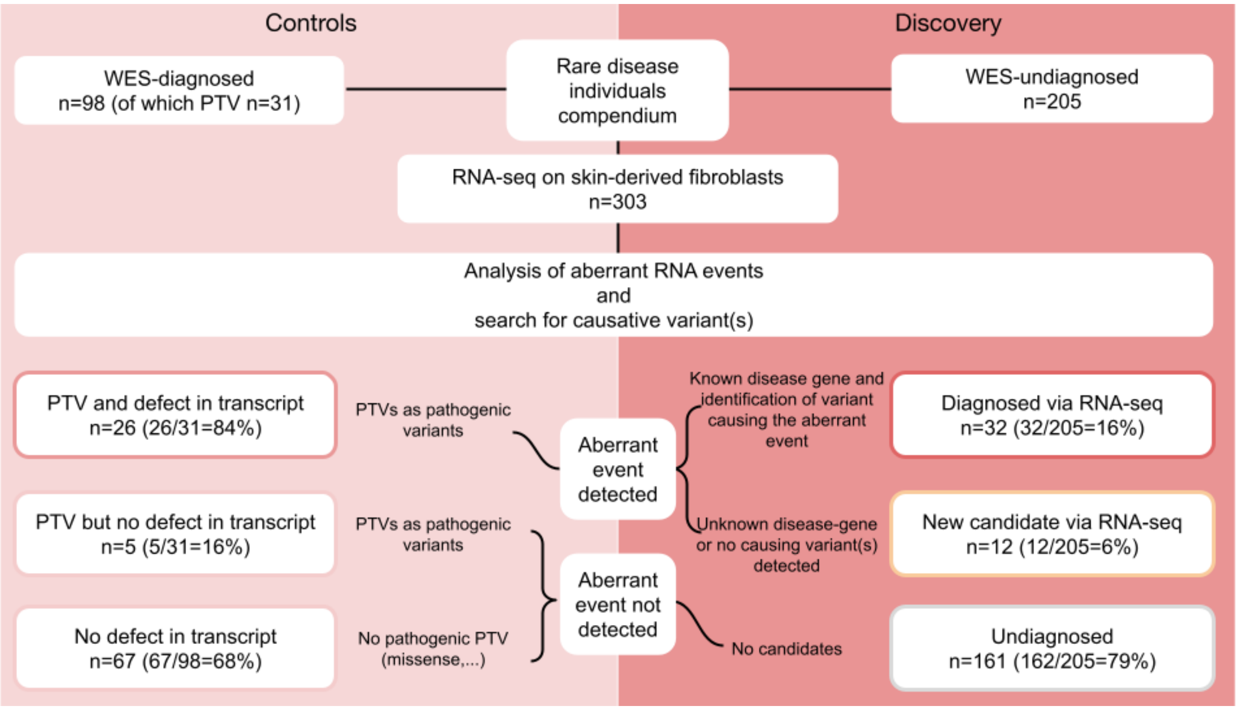


**Supplementary figure 1: Overview of the study.** A total of 303 patients have undergone WES analysis beforehand, which was inconclusive for 205. Systematic detection of aberrant events and consequent analysis led to genetic diagnosis in 32 (16% of the WES-undiagnosed cases) by establishing a genotype-phenotype association, and pinpoint a candidate gene in 11 (5% of the WES-undiagnosed cases), suggesting the discovery of novel disease-genes and more complex pathomechanisms (right). A transcript defect was also detected in 26 out of 31 (84%) of the WES-diagnosed cases carrying pathogenic protein-truncating variants (left). PTV: protein-truncating variant.

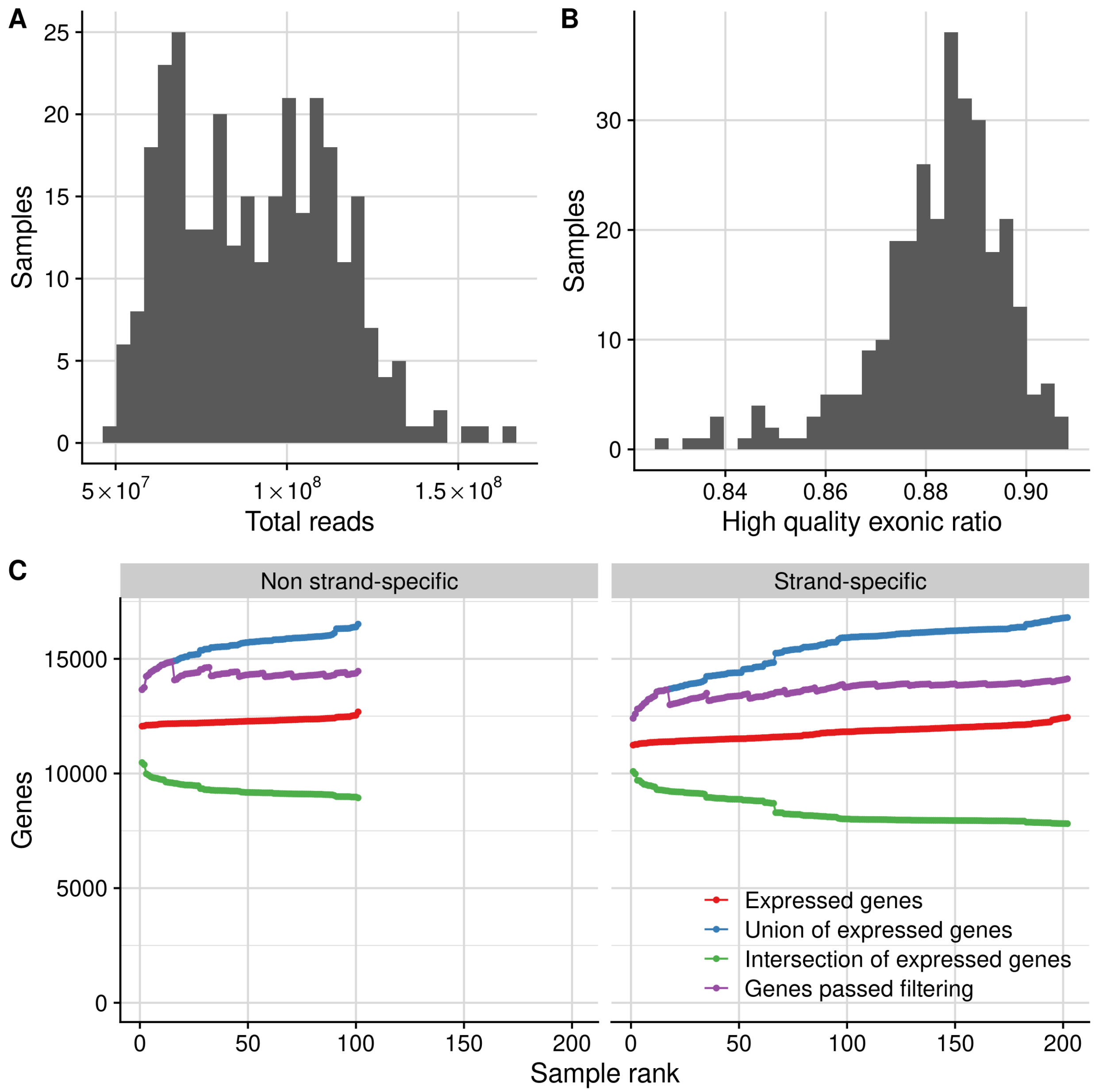


**Supplementary figure 2: Quality control. (A)** Distribution of the total sequencing depth per sample ranging from 50 – 165 million reads. **(B)** Distribution of the high quality exonic ratio per sample, defined as the high quality exonic reads divided by the total high quality reads (exonic + intronic + intergenic). **(C)** Number of expressed genes cumulative across all the samples from the non strand-specific and the strand-specific cohorts. Colors represent the genes expressed on each sample (red), the union of all detected genes (blue), the intersection of expressed genes (green), and the genes that passed the minimum FPKM filter as a group (violet).

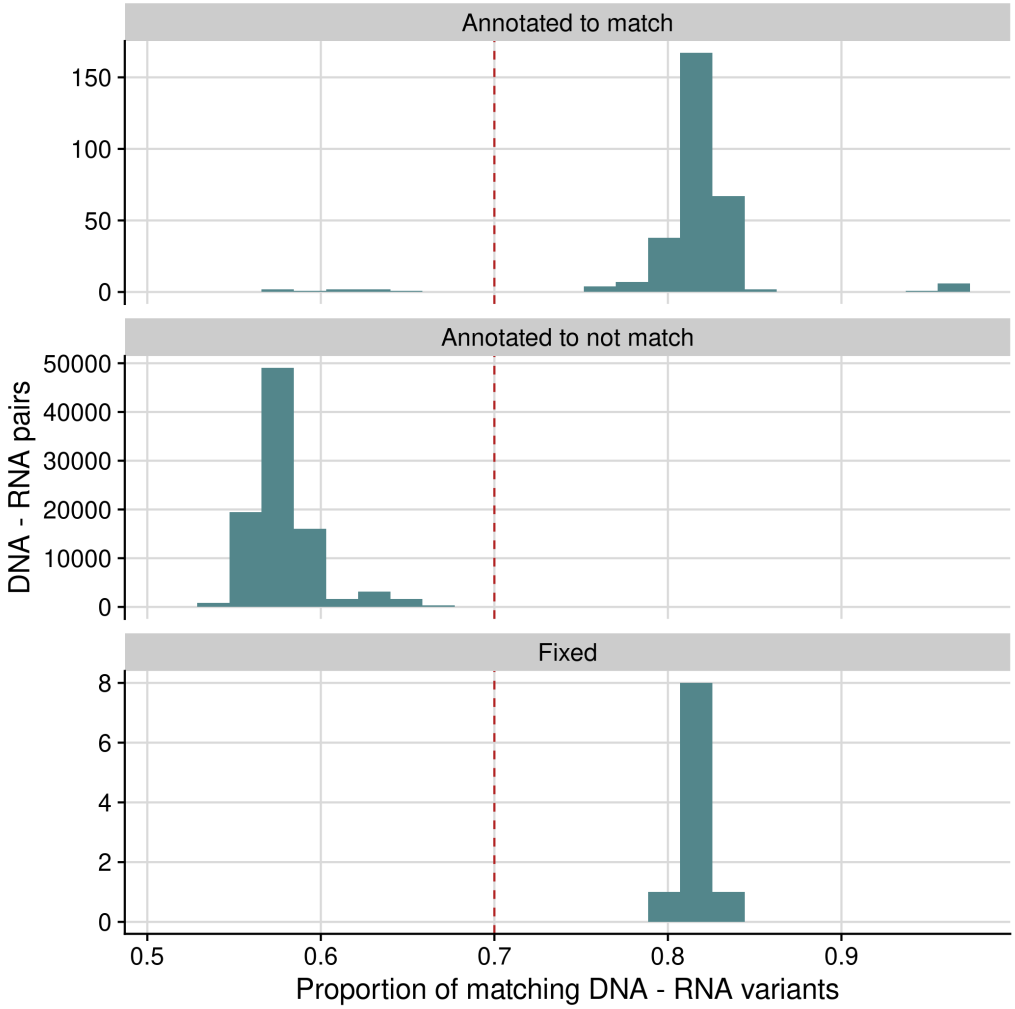


**Supplementary figure 3. DNA-RNA sample matching.** Distribution of the proportion of matching DNA-RNA variants when comparing all sample combinations, stratified by the DNA-RNA sample pairs that were originally annotated to match, to not match, and that were fixed after applying the matching algorithm from DROP (n=10). A cutoff of 0.7 distinctly separated the matching with the non-matching pairs.

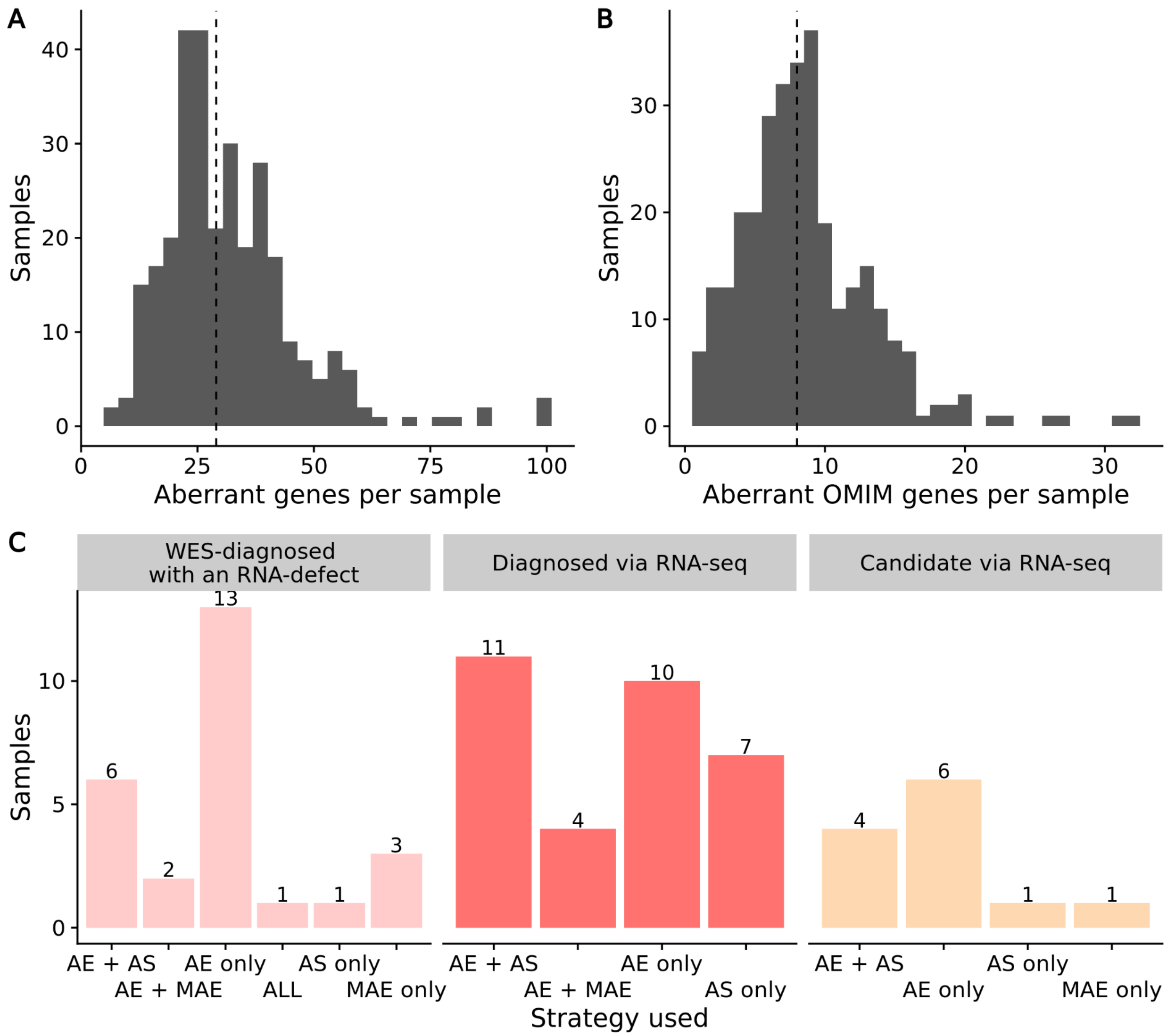


**Supplementary figure 4. Aberrant events per sample. (A)** Distribution of the number of aberrant genes (union of aberrant expression, aberrant splicing, and MAE). Genes harboring many splice aberrations or rare MAE variants are counted only once. The dashed line corresponds to the median at 29. **(B)** Same as (A) but for genes known to cause a Mendelian disease (OMIM genes). The dashed line corresponds to the median at 8. These correspond to the genes of clinical relevance. **(C)** Causative and candidate RNA-seq aberrant events detected by our pipeline across the 26 WES-diagnosed cases with a protein-truncating variant and defect in RNA-seq (Supplementary figure 1), the 32 RNA-seq diagnosed cases, and the 12 RNA-seq candidates cases. AE: aberrant expression, AS: aberrant splicing, MAE: mono-allelic expression.

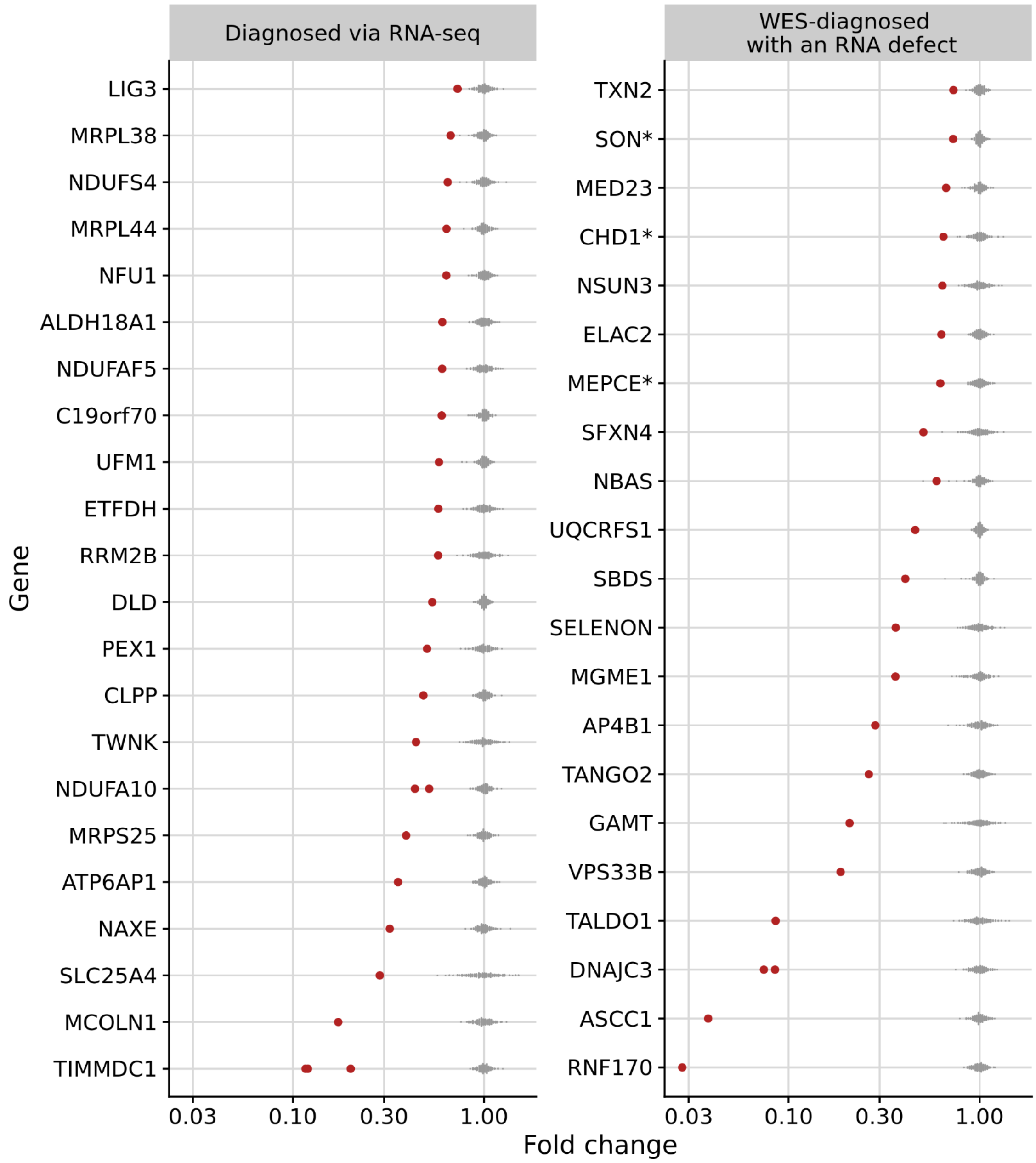


**Supplementary figure 5: Expression fold change of samples with aberrant expression.** Gene expression fold change (normalized counts / mean across samples) of all disease-causal genes that were aberrantly expressed in their corresponding affected sample. Each dot corresponds to a sample, with the affected ones in red. Data stratified by cases diagnosed via RNA-seq (n=25) and diagnosed via WES (n=22). Genes with a dominant mode of inheritance are marked with a *. The two *NDUFA10* cases are siblings, as well as the two *DNAJC3* cases. The three *TIMMDC1* cases are unrelated.

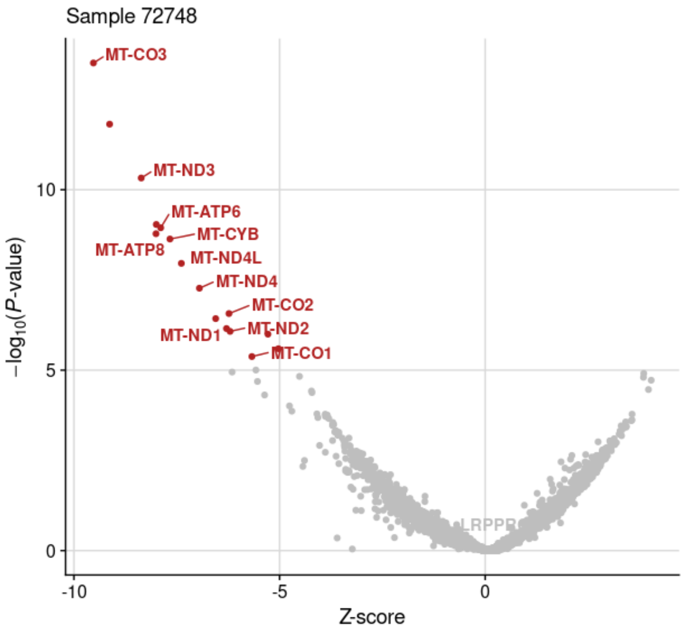


**Supplementary figure 6: Case with many mtDNA expression outliers.** Gene-level significance (-log10(*P*), y-axis) versus Z-score of sample 72478, with 11 mtDNA genes labeled among the 15 underexpression outliers (red dots). The disease causal gene, *LRPPRC*, is involved in the post-transcriptional regulation of mitochondrial gene expression, thus explaining the aberrant mtDNA genes.

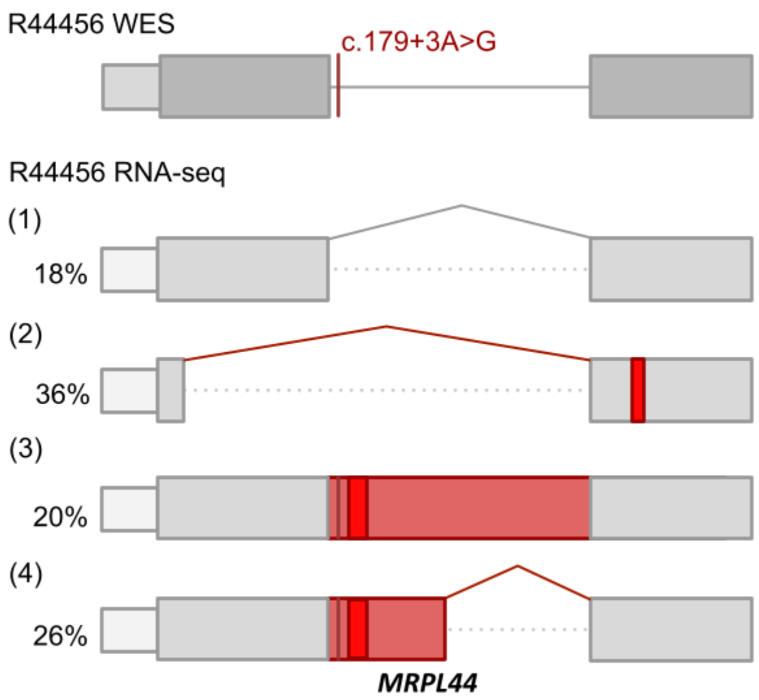


**Supplementary figure 7: Complex pattern of aberrant splicing.** Schematic depiction of the complex pattern of aberrant splicing of *MRPL44* in sample R44456 due to a homozygous splice region variant (c.179+3A>G). The percentage of detected transcript isoform is shown next to it.

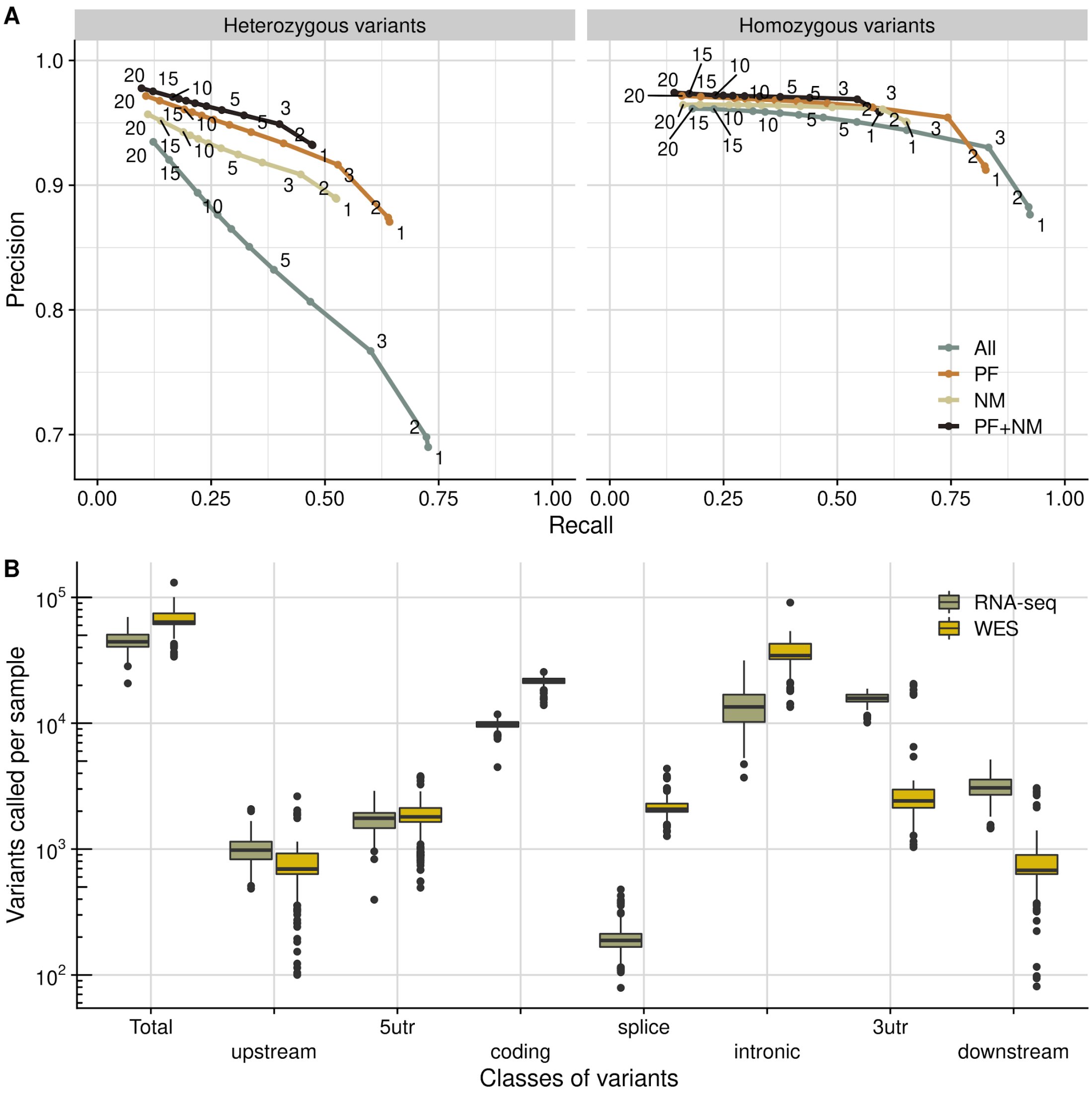


**Supplementary figure 8: Analysis of variants called by RNA-seq. (A)** Average precision (true positives / all positives) vs. average recall (false positives / (false positives + true positives)) of variants detected by RNA-seq in comparison to variants detected by WGS in 210 samples from suprapubic skin from GTEx. The average is taken across samples. Colors indicate all variants called by RNA-seq, the ones that passed the GATK filters (PF), the ones that were not in a masked region (NM), and the combination of both filtering strategies (Materials and Methods). Data points are labeled by the minimum number of reads required to support the alternative allele. This analysis led us to apply the GATK filters and to consider variants not in masked regions and with a minimum alternative allele count of 3, where we see for both homozygous and heterozygous variants a strong precision drop. More stringent restrictions to variant coverage further improve the average precision, which comes at the cost of the recall. **(B)** Distribution of the variants called by WES or RNA-seq (y-axis) in total and stratified by variant classes (x-axis) per sample.

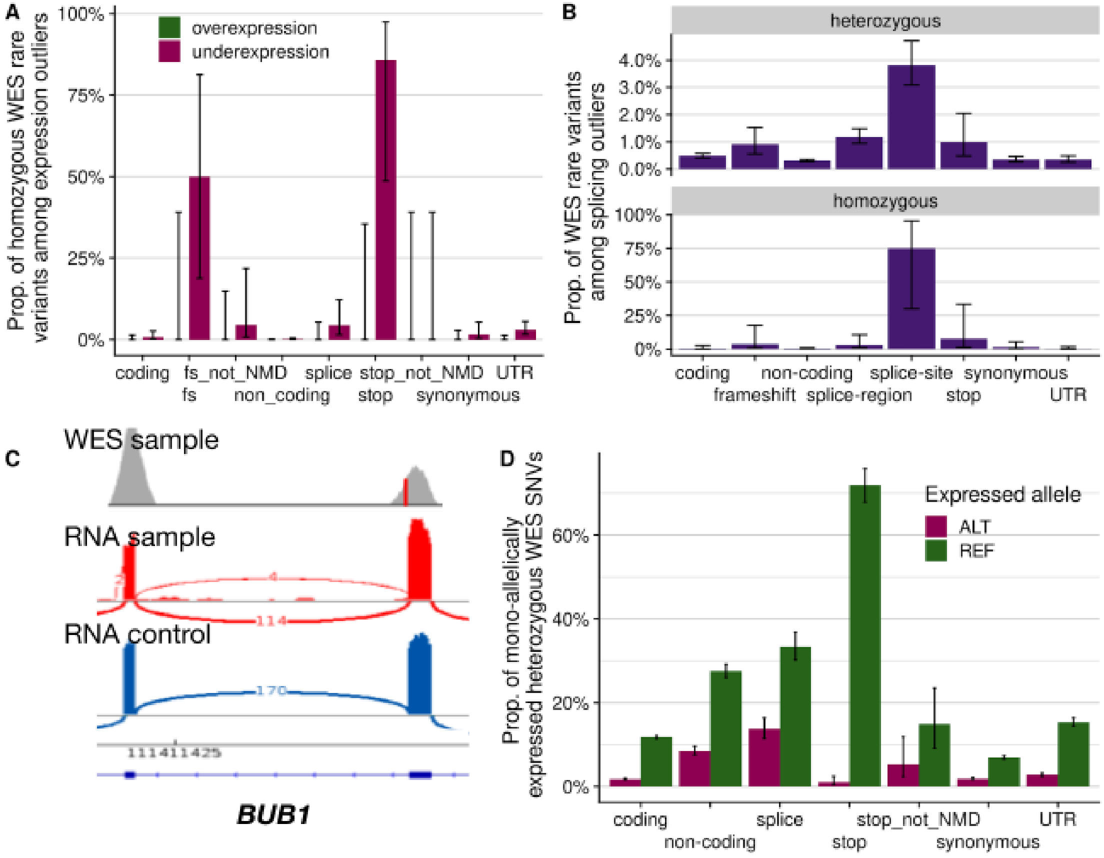


**Supplementary figure 9: Rare variants leading to outliers.** **(A)** Proportion of homozygous WES rare variants stratified by VEP class in genes that are aberrantly expressed. Error bars represent 95% confidence intervals of a binomial test (also in panels (B) and (D)). **(B)** Proportion of WES rare variants (both heterozygous and homozygous) stratified by VEP class in genes with an aberrantly spliced junction. **(C)** Sashimi plot presenting normal splicing on the gene *BUB1* in spite of a homozygous variant in the direct splice-site (shown in red) of a candidate sample. **(D)** Proportion of rare heterozygous WES SNVs that are mono-allelically expressed towards either the reference or alternative allele, stratified by VEP class. In panels (A) and (D), 'not NMD' corresponds to stop or frameshift (fs) variants that are predicted to escape NMD due to being in either the last exon of the gene or in a position that is not expressed in fibroblasts.

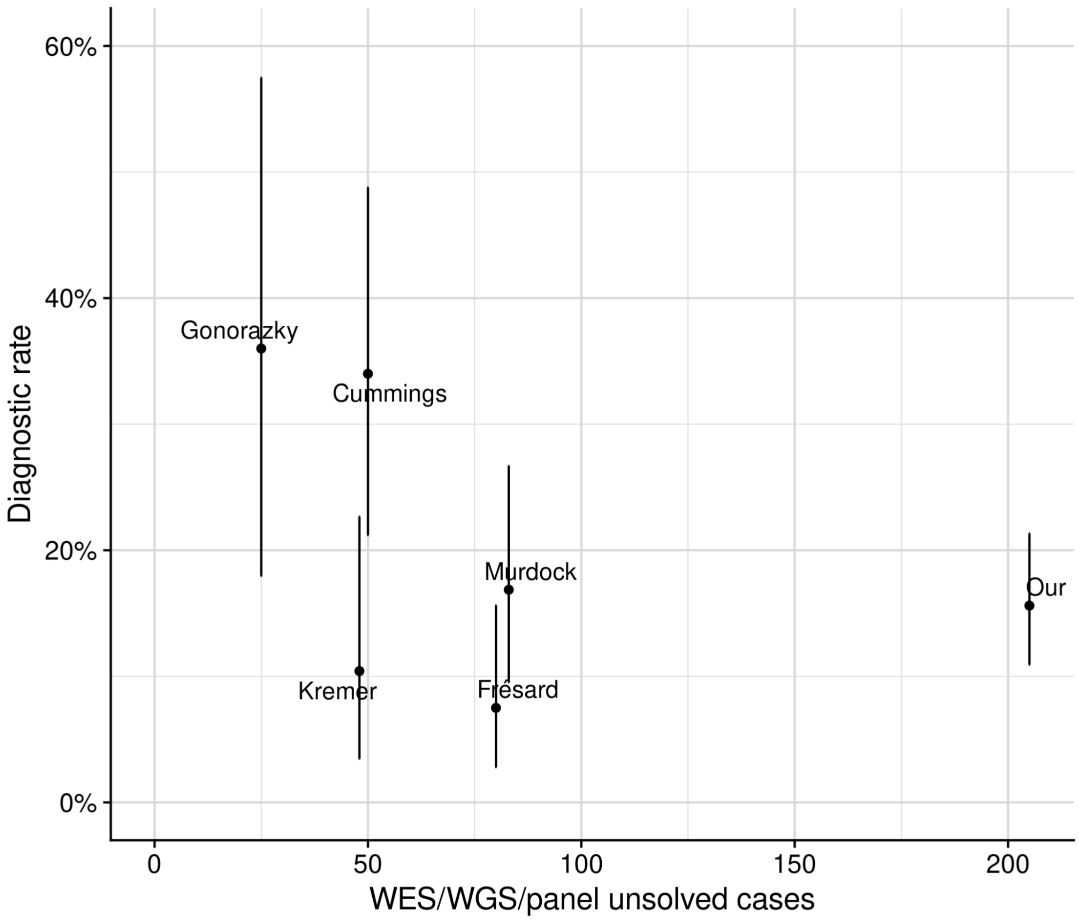


**Supplementary figure 10: Diagnostic rate across different cohorts.** Diagnostic rate (solved cases / cohort size) vs. cohort size, defined as the number of cases that were unsolved after either WES, WGS, or panel sequencing, across five published cohorts (23-26, 28) and our study. Error bars stretch a 95% confidence interval of a binomial test. A diagnostic rate exceeding 30%, such as in Cummings *et al.* (24) and Gonorazky *et al.* (26), could be reached by more stringent inclusion of patients with either variants of uncertain significance in splice regions or unsolved cases where phenotype is a strong predictor of genotype.
